# Cost-effectiveness of immunising interventions to reduce respiratory syncytial virus disease burden in infants in Australia

**DOI:** 10.1101/2025.08.06.25333104

**Authors:** Julian B. Carlin, Adrian J. Marcato, Yingying Wang, Robert Moss, Kylie S. Carville, Xinghui Chen, Victoria L. Oliver, Violeta Spirkoska, Patricia T. Campbell, David J. Price, Natalie Carvalho, Jodie McVernon

## Abstract

**Background:** Two immunising products are emerging to prevent the burden of respiratory syncytial virus (RSV) in infants: long-lasting monoclonal antibodies (mAbs) and maternal vaccines given during pregnancy (MV). This study assesses the potential cost-effectiveness of programs involving each product, to help inform policy decisions related to their implementation in the Australian context.

**Methods:** We developed an individual-based dynamic transmission model of RSV infection, linked to a clinical pathways model and cost-effectiveness model. We modelled key scenarios exploring varying eligibility and coverage of immunisation products for at-risk and not-at-risk populations, in addition to sensitivity analyses of immunisation characteristics, program costs, and the impact of potential under-ascertainment of RSV burden. We estimated the cost-effectiveness of each program from a health system perspective, with results presented as incremental cost-effectiveness ratios in terms of cost per quality-adjusted life year gained (QALY).

**Findings:** We found a combined program in which administration of MV during pregnancy is supplemented with a birth-dose of mAbs for newborns born without protection from MV is likely to be cost-saving, compared to the status quo of no MV or mAbs delivered. This program averted on average 41% of infant hospitalisations per year and reduced QALY losses by 33%.

**Interpretation:** Programs combining infant immunisation products are likely to significantly reduce the burden of RSV disease in Australia, and be cost-saving. However, their estimated impact and cost-effectiveness is strongly dependent on key assumptions i) the consistency and completeness of ascertainment of disease burden over time; ii) the cost of a hospitalisation and immunising dose; iii) the efficacy and durability of protection of the modelled products, and; iv) the timing and coverage of the immunisation delivery.

**Funding:** This modelling was commissioned by the National Immunisation Division of the Australian Government Department of Health, Disability and Ageing.

## 1. Introduction

Respiratory syncytial virus (RSV) causes a range of respiratory illnesses in infants and young children. RSV can cause mild upper respiratory tract infections, including the common cold and otitis media, and more severe lower respiratory tract infections such as bronchiolitis and pneumonia requiring hospitalisation. Additionally, RSV infection in early childhood has been associated with an increased risk of developing asthma later in life [1].

There is a high global burden of RSV disease in children less than five years of age, with infants <6 months of age experiencing the highest burden of hospitalisations in 2019 [2]. This trend is also observed in Australia, with children less than five years of age comprising approximately 75% of all RSV-coded hospitalisations in 2016–2019 [3]. In particular, very young infants <2 months of age experience the highest burden of RSV disease, with hospitalisation incidence rates of 7,200 per 100,000 [3].

Until recently, the short-acting monoclonal antibody product Palivizumab (Synagis), was the only prophylactic intervention available globally to reduce the risk of RSV in infants. It is administered to at-risk infants (e.g. premature infants and infants with congenital heart disease) as a monthly intramuscular injection during the RSV season, which typically precedes the influenza season and runs for several months across autumn and winter. However, the impact of this product is limited as protection from one dose is short-lived (approximately one month), and the requirement for several doses increases the cost and complicates the implementation of associated immunisation programs.

New options are now available for the prevention of RSV-associated severe respiratory disease in infants. These products involve passive immunisation of infants through receipt of one dose of RSV-specific longacting monoclonal antibodies (Nirsevimab, Beyfortus) or antenatal vaccination of pregnant people (Abrysvo) [4–6]. The population-level impact and cost-effectiveness of immunisation programs using these approaches is a current concern for policy makers as these products are near to market [7, 8].

To explore the potential impact and cost-effectiveness of these new products, we developed an individual-based model of RSV infection and disease that we report here as a case study using Australian data. Our modelling framework, designed in close consultation with the Australian Technical Advisory Group on Immunisation (ATAGI) and the Australian Government Department of Health, Disability and Ageing, was one piece of evidence that informed National Immunisation Policy (NIP) decisions in 2024. In this paper, we analyse and report on the anticipated health system impact and cost-effectiveness of feasible immunisation programs using Nirsevimab (henceforth referred to as mAbs), maternal vaccination with Abrysvo (henceforth referred to as MV), and combinations thereof, to reduce the burden of RSV disease in Australian infants <1 year of age.

## 2. Methods

We estimate the cost-effectiveness of different programs of immunisation products, via a pipeline of three distinct models. These models, as illustrated in Figure 1, are i) a transmission model of epidemic spread and immunisation product roll-out, ii) a clinical pathways model that probabilistically simulates clinical endpoints given an infected individual’s age, risk status, and immunisation status, and iii) a cost-effectiveness model that computes incremental costs and quality-adjusted life-years (QALYs) compared to the no-intervention scenario.

**Figure 1.**
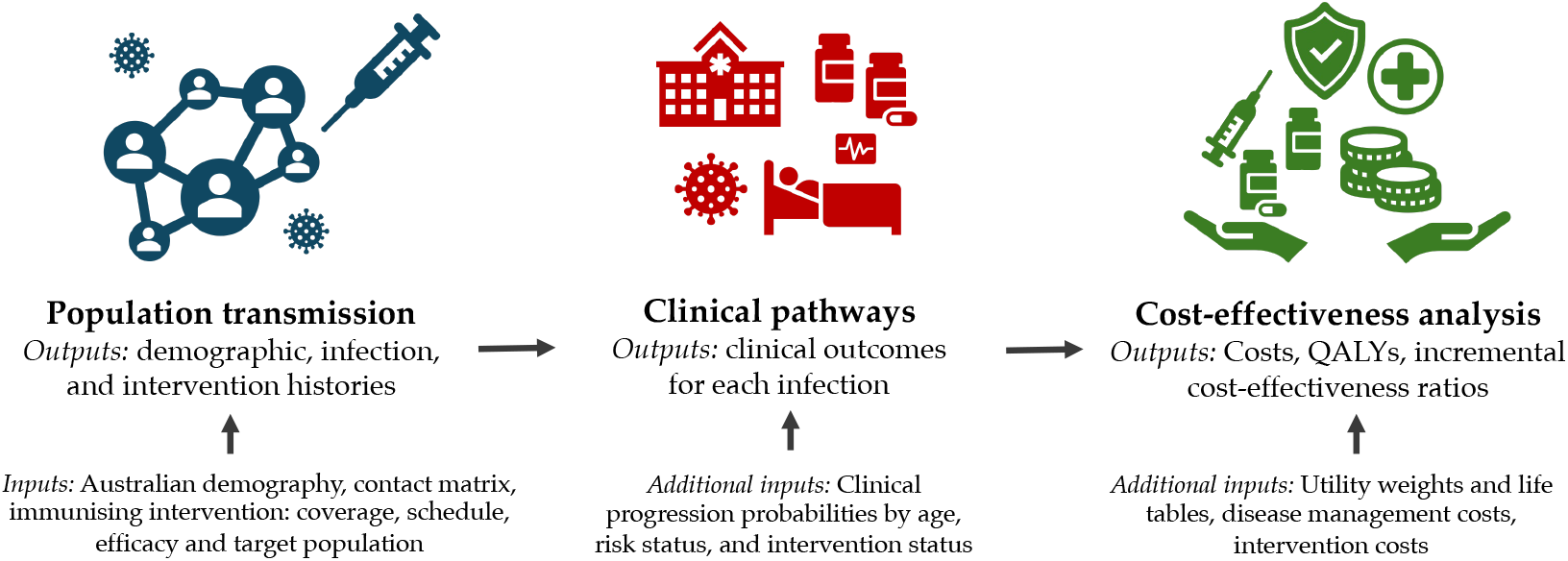
Schematic overview of the inputs and outputs of the transmission, clinical pathways, and cost-effectiveness models.

### 2.1. Infection and transmission model overview

The goal of the population transmission model is to reproduce key trends of RSV infection in Australia, and to project likely transmission dynamics given introduction of different combinations of immunisation products.

The transmission model is a dynamic individual-based model. We track the age (to the granularity of days), sex, age at death, and the demographic status of each individual. We explicitly model pregnancy, and connect infants and pregnant people. We matched to Australian data for the age-sex distribution, age- and sex-specific mortality rates, fertility rates, and the gestational age of newborns upon birth. The simulated population of 100,000 individuals includes a birth cohort per year of roughly 1,300 infants, matching Australian demographic data. We assumed age-stratified population-level mixing of individuals as estimated for the Australian population by Mistry et al. [9]. Details on demographic data sources and simplifying choices are outlined in Appendix A in the supplementary materials.

The model assumes that repeated exposures over the life course of an individual build natural immunity to infection with RSV. We do so via a tiered susceptible-exposed-infectious-recovered-susceptible (SEIRS) model of RSV infection, where after recovery following infection, subsequent infection is less likely. This compartmental structure is commonly used for RSV models [10, 11]. Further mathematical details of the transmission model are in Appendix B in the supplementary materials.

Given limited Australian data on the incidence of RSV in settings other than hospitals, we calibrated the uncertain parameters of the transmission model and the probability of hospitalisation given infection to historical RSV-coded hospitalisation data (2018–2019) provided by the Department of Health, Disability and Ageing using Admitted Patient Care data supplied by the Australian Institute of Health and Welfare (AIHW). We also ensured the infection dynamics were consistent with the accepted heuristic that 75% of infants are infected with RSV in their first year of life, and 95% by two years of age [12, 13]. We used hospitalisation data from 2018–2019 given clear COVID-19 impacts on RSV circulation in Australia from 2020–2022, and because data were not available beyond 2022. The data sources and further details of model calibration are in Appendix C in the supplementary materials.

### 2.2. Clinical pathways model overview

The model has six mutually exclusive clinical endpoints for each RSV infection that are directly relevant to healthcare costs. As illustrated in Figure 2, these endpoints are: no medical care is sought, a general practitioner visit (GP), emergency department consultation with no admission to a hospital ward (ED), admission to a hospital ward (Hospitalisation) without further progression, admission to an intensive care unit (ICU), and death following admission to an ICU (Death).

**Figure 2.**
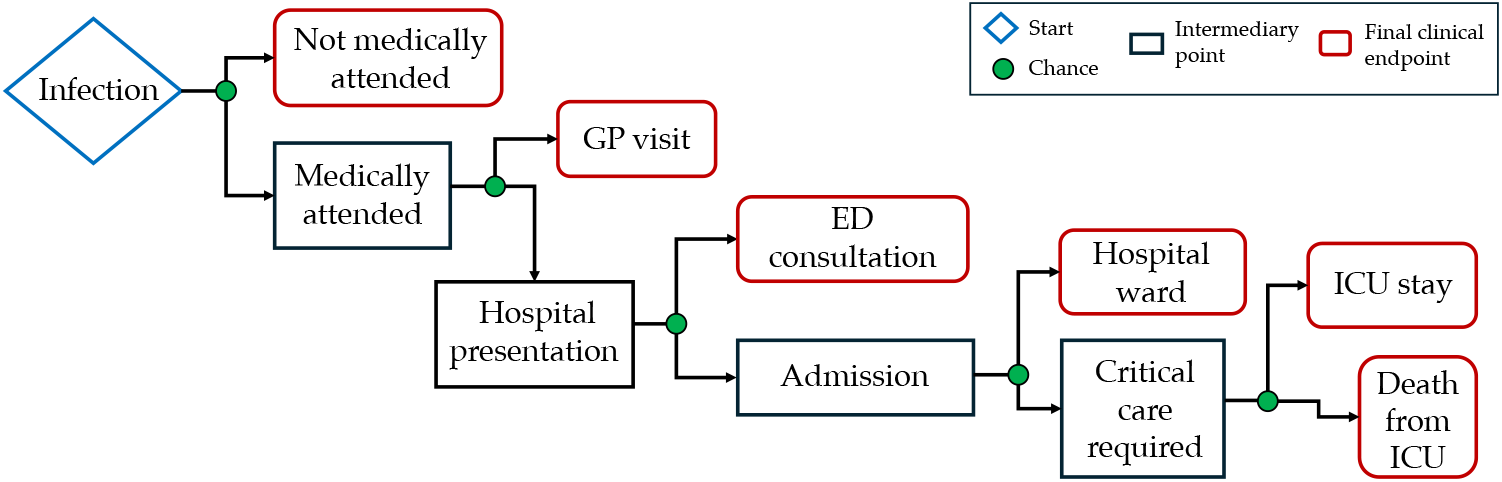
Flowchart illustrating the clinical pathways model.

We mapped infections to clinical endpoints in an age- and risk-specific manner, using Australian data wherever possible. We probabilistically assigned only the final clinical outcome experienced for each RSV infection, assuming that medically attended RSV infections progress sequentially through the clinical pathway. For example, we assumed that an individual who is assigned a clinical endpoint of ICU, previously presented to the emergency department and was admitted to a hospital ward. Similarly, we assumed that fatalities only occur following admission to ICU. Given limited population uptake we did not explicitly assume any impact of short-acting monoclonal antibodies (Palivizumab) in the population-level data to which we calibrated the clinical pathways model.

Premature infants born prior to 37 weeks’ gestational age were assumed to have a higher risk of severe outcomes (“at-risk”) than infants born after 37 weeks of gestation (“not at-risk”). We used line-listed data (2017–2019) from a large, tertiary paediatric hospital in Melbourne to estimate the relative risk (*RR*) for hospitalisation and ICU admission (for an at-risk infant compared to a not at-risk infant) as 2.46 and 5.69, respectively. These data were selected to align temporally with our national hospital admission data (2018– 2019) as closely as possible. These estimates align broadly with those reported from another Australian paediatric hospital (2015–2022) [14]. In the absence of data from other clinical settings, we applied the *RR* for hospitalisation to general practice and emergency department visits, and the *RR* for ICU admission to fatality. Given limited local evidence on the prevalence and relative risk of RSV outcomes for underlying clinical conditions other than prematurity, no other at-risk conditions were explicitly included in our model.

Further details of the clinical pathways model, including data sources and assumptions are in Appendix D in the supplementary materials.

### 2.3. Immunisation efficacy

Evidence from clinical trials and early post-licensure observational studies demonstrate that both mAbs and MV prevent severe medically-attended disease outcomes in infants [5, 6, 15–19, 4]. However, the reported clinical efficacy and effectiveness endpoint estimates from studies of each product (e.g. efficacy against lower respiratory tract disease with two or three symptoms) do not map directly to our endpoints of interest, which instead relate to healthcare utilisation given RSV infection. Considering all available evidence we determined in consultation with ATAGI a set of immunisation efficacy values that are consistent with trial results to use as input values in the model for both MV and mAbs, and for not at-risk and at-risk infants. These mappings resulted in the initial efficacy for each endpoint listed in Table 1. Given the absence of evidence on immunisation efficacy of both products against infection, we ascribed minimal efficacy for this endpoint.

**Table 1:** Initial efficacy against infection and clinical endpoints for mAbs and MV, for both at-risk and not-at-risk infants.

|  |  | Infection (%) | GP (%) | ED (%) | Hosp. (%) | ICU (%) | Death (%) |
| --- | --- | --- | --- | --- | --- | --- | --- |
| mAbs | Not at-risk | 10 | 50 | 70 | 80 | 85 | 100 |
|  | At-risk | 10 | 45 | 65 | 75 | 80 | 100 |
| MV | Not at-risk | 6 | 30 | 50 | 75 | 85 | 100 |
|  | At-risk | 6 | 25 | 50 | 70 | 80 | 100 |

Abbreviations: mAbs = monoclonal antibodies; MV = maternal vaccination; GP = general practitioner visits; ED = emergency department (non-admitted) visits; Hosp. = hospitalisations; ICU = intensive care unit admissions.

In each scenario we model, mAbs are only given to newborns and MV is provided to pregnant people between 28 and 36 weeks gestation. The benefits of MV are only considered conferred to newborns born at least two weeks after the vaccination. We assume that the mAbs immunisation efficacy is at a maximum for 90 days, and the protection wanes exponentially thereafter, with a half-life of 70 days from that point [6, 20]. For MV, we assume the immunisation efficacy is at a maximum for 90 days post-vaccination, and the protection wanes linearly to zero in the 180 days hence [4]. In the model, the complement of the product specific efficacy against each endpoint at the time of an individual’s infection is multiplied by their age- and risk-specific probability of that endpoint (see Section 2.2), to further reduce the probability of each clinical endpoint. These assumptions are further outlined in Appendix D in the supplementary materials.

### 2.4. Scenarios

We model the impact of immunisation programs involving only mAbs (single dose), only MV, and programs involving combinations of mAbs and MV.

We assume mAbs are only delivered to newborns for two reasons: i) the bulk of the disease burden is seen in infants under four months of age, and ii) given logistical and programmatic considerations that real-world implementation is most likely to be in a hospital setting prior to discharge after birth of the infant, as part of a suite of products given to infants. For mAbs-only programs, we explore the impact of varying eligibility by at-risk status, as well as the timing and duration of delivery and uptake coverage. We consider both a year-round and seasonal program, and consider coverage rates of 50% and 70%. A “seasonal” program was defined as February through July to maximise the potential impact, without compromising the efficiency of the overall program. A seasonal program over these months helps to ensure the bulk of the uptake of mAbs (when they are at their highest efficacy) is just prior to and during a typical RSV season. We explore alternative durations and timings for seasonal delivery in Appendix E in the supplementary materials.

For MV immunisation programs, we only explore the impact of varied coverage levels. We do not consider seasonal MV programs, in order to align with current year-round maternal vaccine programs for other diseases in Australia [21].

Combinations of mAbs and MV are also explored with varied coverage and eligibility. These programs were explored as likely “feasible” programs, that aimed to maximise protection of infants while minimising the use of the more-expensive mAbs (compared to MV). Combination scenarios explored the likely benefit of selectively administering mAbs to newborns not considered protected from MV (based on the date of MV delivery and/or gestational age).

Each program’s impact on clinical endpoints is assessed over a two-year time horizon, by comparing to a no-intervention scenario, i.e. a scenario without delivery of either immunisation product.

### 2.5. Cost-effectiveness model overview

A cost-utility analysis was conducted to evaluate the cost-effectiveness of each program from a healthcare system perspective following national guidelines [22]. Note that this analysis was performed independently of national cost-effectiveness analysis conducted by the Pharmaceutical Benefits Advisory Committee (PBAC). We present results as an incremental cost-effectiveness ratio (ICER), representing the incremental costs per QALY gained compared to no intervention. The economic model uses outputs from the clinical pathways model described above, ensuring consistency in the relevant demographics, infection history, immunisation characteristics, and infection-related outcomes.

The economic analysis only considered medically-attended clinical outcomes in infants over the two-year time horizon, with QALY losses due to premature mortality captured over a lifetime. QALYs gained were estimated as detailed in Appendix F.2 in the supplementary materials. Briefly, health related quality of life decrements for each clinical outcome were sourced from Shoukat et al. [23] and the duration of each health state was based on length of stay data from the National Hospital Cost Data Collection (NHCDC) dataset [24]. Programmatic costs (dose and administration costs) and costs of healthcare for medically-attended cases were included. The dose prices for MV and mAbs in Australia are unknown. We assumed prices of A$290 for mAbs and A$160 for MV in the primary analyses based on United States pricing [25], scaled down by 43.6% to reflect reported average difference in medicines prices between the United States and Australia [26]. These estimated prices were varied in sensitivity analyses based on prices reported in the literature in other settings, or used in cost-effectiveness analyses in Australia and elsewhere. A wastage rate of 5% was assumed for both. The cost of dose administration and outpatient visits for non-admitted cases requiring a GP consultation were based on Medicare Benefits Schedule pricing. The cost of outpatient ED visits and hospitalisations were based on the NHCDC dataset. The full details of how costs were estimated are included in Appendix F.3 in the supplementary materials.

All costs are reported in 2024 Australian dollars (A$). All future costs and QALYs are discounted at 5% per annum in line with national guidelines [22], with the discount rate varied in the sensitivity analysis. Economic evaluation methods are described in detail in Appendix F and the analysis is conducted and reported in accordance with the Consolidated Health Economic Evaluation Reporting Standards (CHEERS) 2022 [27].

### 2.6. Sensitivity analyses

We performed deterministic – one-way and two-way – and probabilistic sensitivity analyses to explore the impact of uncertainty in key model parameters, including immunisation efficacy, potential bias in the ascertainment of hospitalisations and other clinical endpoints, immunisation product costs and healthcare resource costs, and changes in the discount rate. We detail the parameter ranges and distributions explored in these analyses in Appendix H in the supplementary materials.

### 2.7. Ethical approval

This study was funded by the National Immunisation Division of the Australian Government Department of Health, Disability and Ageing to support policy decision making. The model was parameterised using nationally aggregated patient data provided by the Department of Health, Disability and Ageing using Admitted Patient Care data supplied by the Australian Institute of Health and Welfare (AIHW). Approval for the use of these data for analyses and subsequent publication was granted by the Department. Clinical risk factors in early infancy were parameterised using de-identified patient data from the Royal Children’s Hospital, use of which was granted by the Murdoch Children’s Research Institute Ethics Committee (#37185)

### 2.8. Software used

The transmission and clinical pathways models were written in `Python`, utilising the `Polars` dataframe library. The cost-effectiveness model was written in `R` (version 4.3.3) and the economic analysis was primarily performed using `Tidyverse` package. The package details and code are available at https://gitlab.unimelb.edu.au/julian.carlin/infant-rsv-modelling.

### 2.9. Role of the funding source

This work was funded by the National Immunisation Division of the Australian Government Department of Health, Disability and Ageing. The funders played a role in i) designing the research question, in line with the relevant policy question, ii) facilitating access to data extracts and input from relevant technical advisers (e.g. ATAGI); and iii) granting approval to publish the manuscript. However, the funders did not play a role in the development of the model or the interpretation of the results.

## 3. Results

The dynamic transmission model was calibrated to Australian data (RSV-coded hospitalisations) from 2018– 2019. We see in Figure 3 that the model recovers the observed monthly hospitalisations per 100,000 in month-of-age age-groups. We also verify that the average number of yearly ICU admissions and deaths in the model are consistent with 2018–2019 Australian data. The procedure and details of calibration are included in Appendix C in the supplementary materials.

**Figure 3.**
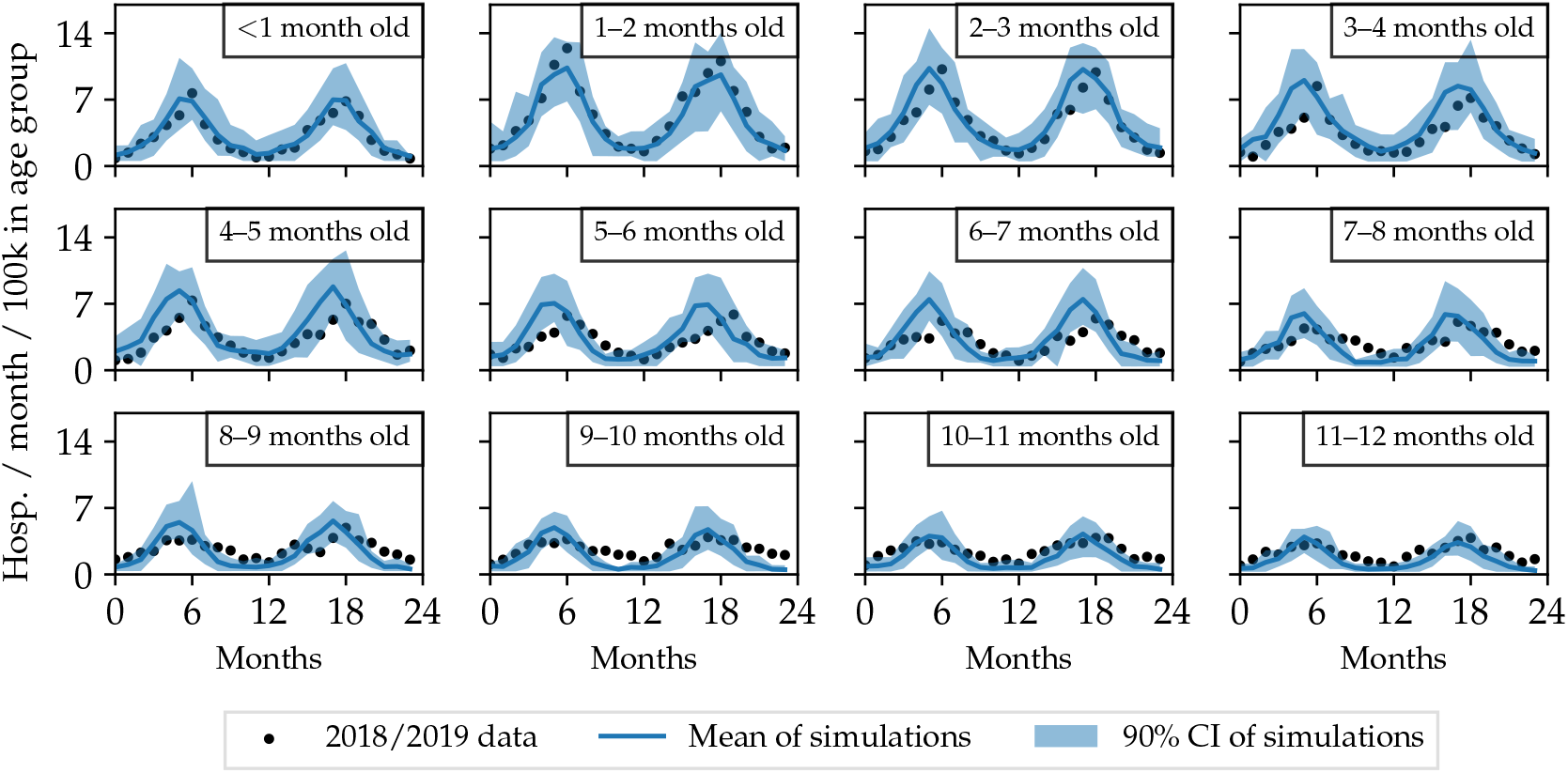
Monthly RSV-coded hospitalisations per 100,000 individuals in each age group generated from the transmission and clinical pathways model (blue curve is the mean, blue band is the 5th–95th percentile from 20 transmission and 1000 clinical pathways simulations), compared to observed monthly hospitalisations in 2018/2019 (black points).

Figure 4 shows the average modelled annual incidence of RSV infections, GP and ED visits, hospitalisations, ICU admissions, and deaths in infants for four exemplar scenarios: no intervention, mAbs given to 50% of newborns year-round, MV given to 70% of pregnant people year-round, and a combination scenario in which 70% of pregnant people are given MV year-round, and 50% of newborns born without the protection of MV are given mAbs year-round. We see that the scenario combining mAbs and MV reduced the incidence of severe clinical outcomes more than only MV or only mAbs (although note that different numbers of doses are delivered in each scenario shown).

**Figure 4.**
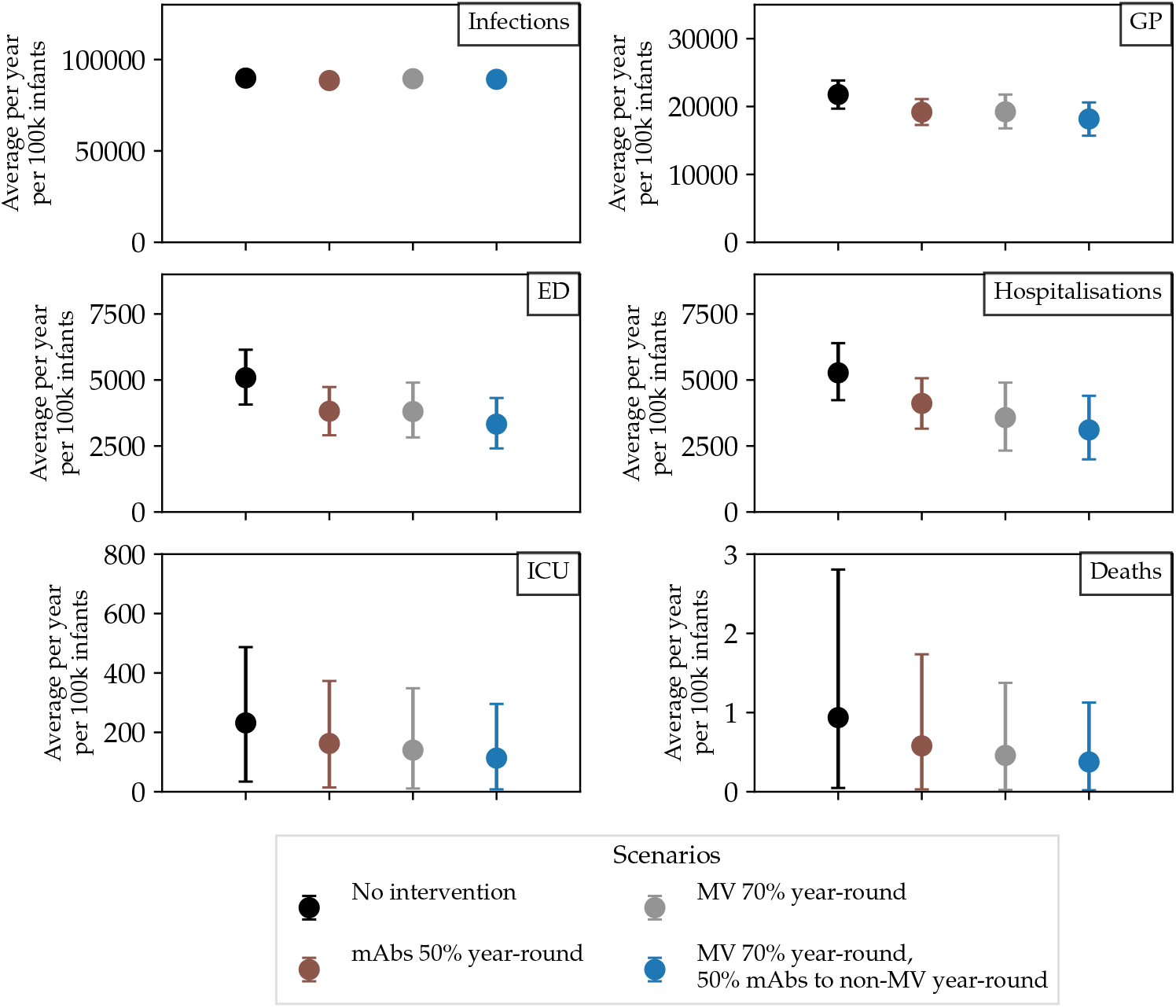
Average incidence (5th–95th percentile) of medically-attended RSV (per 100,000 infants <1 year of age) that leads to GP, ED visits, hospitalisations, ICU admissions and deaths in the no-intervention scenario, a scenario with only mAbs, a scenario with only MV, and a scenario combining mAbs and MV. Abbreviations: GP = general practitioner visits; ED = emergency department (non-admitted) visits; ICU = intensive care unit admissions; MV = maternal vaccination.

For mAbs-only scenarios, the scenario that prevented the most clinical outcomes is when all infants are given mAbs at birth, as expected. We find that similar numbers of clinical outcomes are averted when providing mAbs to 70% of newborns year-round compared to all newborns seasonally (six months, February through July), though the former requires 40% more doses. For MV-only scenarios, we find that the number of outcomes averted increases linearly with the uptake given minimal protection against infection conferred by the immunising products (see Table 1). That is, we do not see any indirect effects. Detailed results for scenarios involving only mAbs and only MV scenarios are included in Appendix G in the supplementary materials.

The numbers of averted cases of each clinical endpoint, with the average doses of MV and mAbs delivered under each scenario which combined the use of mAbs and MV, are tabulated in Table 2. All scenarios lead to a significant reduction in disease burden compared to the no-intervention scenario. The combination scenario with the greatest reduction in disease burden is a year-round program with 70% of pregnant people between 28–36 weeks gestation given MV, in addition to a birth-dose of mAbs for 50% of newborns born without protection from MV, noting the higher average number of mAbs delivered, as compared to the other combination scenarios.

**Table 2:** Average modelled numbers (5th–95th percentile) of clinical endpoints of RSV infection per year in infants scaled to the size of the Australian population in 2018/2019, in a no-intervention scenario. Also tabulated are the average doses delivered per year and numbers of clinical endpoints that are averted per year under different MV and mAbs combination scenarios.

| **Scenario** |  | **GP**<br>**(’000)** | **ED**<br>**(’000)** | **Hosp.**<br>**(’000)** | **ICU**<br>**(’000)** | **Deaths** |
| --- | --- | --- | --- | --- | --- | --- |
| No intervention |  | 66.0<br>(61–70) | 15.4<br>(13–18) | 16.0<br>(14–18) | 0.7<br>(0–1) | 2.8<br>(0–9) |
|  | **MV/mAbs**<br>**delivered**<br>**(’000)** | **GP**<br>**averted**<br>**(’000)** | **ED**<br>**averted**<br>**(’000)** | **Hosp.**<br>**averted**<br>**(’000)** | **ICU**<br>**averted**<br>**(’000)** | **Deaths**<br>**averted** |
| MV 70% year-round<br>50% mAbs to non-MV<br>year-round | 228/62 | 10.9<br>(6–16) | 5.3<br>(3–8) | 6.6<br>(5–9) | 0.4<br>(0–1) | 1.7<br>(0–5) |
| MV 70% year-round<br>50% mAbs to non-MV<br>seasonal | 227/30 | 9.9<br>(5–15) | 4.9<br>(2–8) | 6.1<br>(4–8) | 0.3<br>(0–1) | 1.6<br>(0–5) |
| MV 70% year-round<br>50% mAbs to at-risk<br>non-MV year-round | 228/8 | 8.8<br>(4–13) | 4.3<br>(2–7) | 5.5<br>(3–8) | 0.3<br>(0–1) | 1.7<br>(0–5) |

Abbreviations: GP = general practitioner visit; ED = emergency department (non-admitted) visits; Hosp. = hospitalisations; ICU = intensive care unit admissions; mAbs = monoclonal antibodies; MV = maternal vaccination.

Table 3 presents the number needed to immunise to avert one RSV-associated medically-attended outcome by type of outcome and immunisation program. More doses are needed to prevent rarer outcomes of ICU or death, compared to GP, ED, and hospitalisation, as expected. When mAbs are delivered seasonally rather than year-round, the efficiency is improved such that, on average, one hospitalisation is prevented for every 35 doses, rather than every 48 doses. Even fewer mAbs doses, on average 23, are needed to prevent a hospitalisation in an at-risk infant, even when the doses are delivered year-round. The number needed to immunise for MV-only and combination MV/mAbs scenarios is similar, requiring slightly fewer doses in total.

**Table 3:** Number needed to immunise to prevent one RSV-associated outcome, based on mean outputs from the clinical pathways model.

| Scenario | GP | ED | Hosp. | ICU | Death |
| --- | --- | --- | --- | --- | --- |
| **mAbs-only** |  |  |  |  |  |
| 100% mAbs year-round | 22 | 45 | 48 | 812 | 169,184 |
| 100% at-risk mAbs year-round | 11 | 21 | 23 | 199 | 42,289 |
| 100% mAbs seasonal | 16 | 31 | 35 | 565 | 114,310 |
| 70% mAbs year-round | 21 | 44 | 48 | 802 | 154,172 |
| 50% mAbs year-round | 21 | 43 | 47 | 787 | 152,581 |
| **MV-only** |  |  |  |  |  |
| MV 100% year-round | 29 | 58 | 44 | 828 | 174,162 |
| MV 70% year-round | 29 | 59 | 44 | 817 | 157,083 |
| MV 50% year-round | 30 | 59 | 45 | 839 | 165,212 |
| **MV and mAbs combination** |  |  |  |  |  |
| MV 70% year-round,<br>50% mAbs to non-MV year-round | 27 | 54 | 44 | 806 | 170,333 |
| MV 70% year-round,<br>50% mAbs to non-MV seasonal | 26 | 52 | 42 | 771 | 157,446 |
| MV 70% year-round,<br>50% mAbs to at-risk non-MV year-round | 27 | 55 | 43 | 754 | 141,633 |

Abbreviations: GP = general practitioner visit; ED = emergency department (non-admitted) visits; Hosp. = hospitalisations; ICU = intensive care unit admissions; mAbs = monoclonal antibodies; MV = maternal vaccination.

Figure 5 shows cost-effectiveness results for five scenarios, including MV-only, mAbs-only and combination vaccination programs. All scenarios shown are cost-saving except where mAbs is given year-round to 50% of newborns. A program with 70% coverage of MV and mAbs given to 50% of at-risk newborns born without protection from MV (both delivered year-round) results in the largest cost-savings (A$109,873 per 100,000 Australian population). The three combination vaccination programs delivered the largest QALY gains among the scenarios shown, with the largest gains observed in a program of MV delivered year-round to 70% of pregnant people, with mAbs given year-round to 50% of newborns born without protection from MV, regardless of risk status. See Table 4 for a summary of cost-effectiveness results for these combination scenarios. See Appendix G in the supplementary materials for cost-effectiveness results for scenarios that consider immunisation with only mAbs or only MV.

**Table 4:** Mean costs, quality-adjusted life years gained, and incremental cost-effectiveness ratios, per 100,000 population (MV and mAbs combination scenarios)

| Scenario | Total Cost | QALY decrements | Incr. Cost | Incr. QALY | ICER (/QALY gained) |
| --- | --- | --- | --- | --- | --- |
| No intervention | A\$1,443,902 | -2.52 | Reference | Reference | Reference |
| MV 70% year-round,<br>50% mAbs to non-MV<br>year-round | A\$1,385,917 | -1.69 | -A\$57,986 | 0.83 | Cost-saving |
| MV 70% year-round,<br>50% mAbs to non-MV<br>seasonal | A\$1,345,192 | -1.74 | -A\$98,711 | 0.77 | Cost-saving |
| MV 70% year-round,<br>50% mAbs to at-risk<br>non-MV year-round | A\$1,334,029 | -1.79 | -A\$109,873 | 0.72 | Cost-saving |

Abbreviations: QALY = quality-adjusted life years; Incr. = incremental; ICER = incremental cost-effectiveness ratio; MV = maternal vaccination; mAbs = monoclonal antibodies.

**Figure 5.**
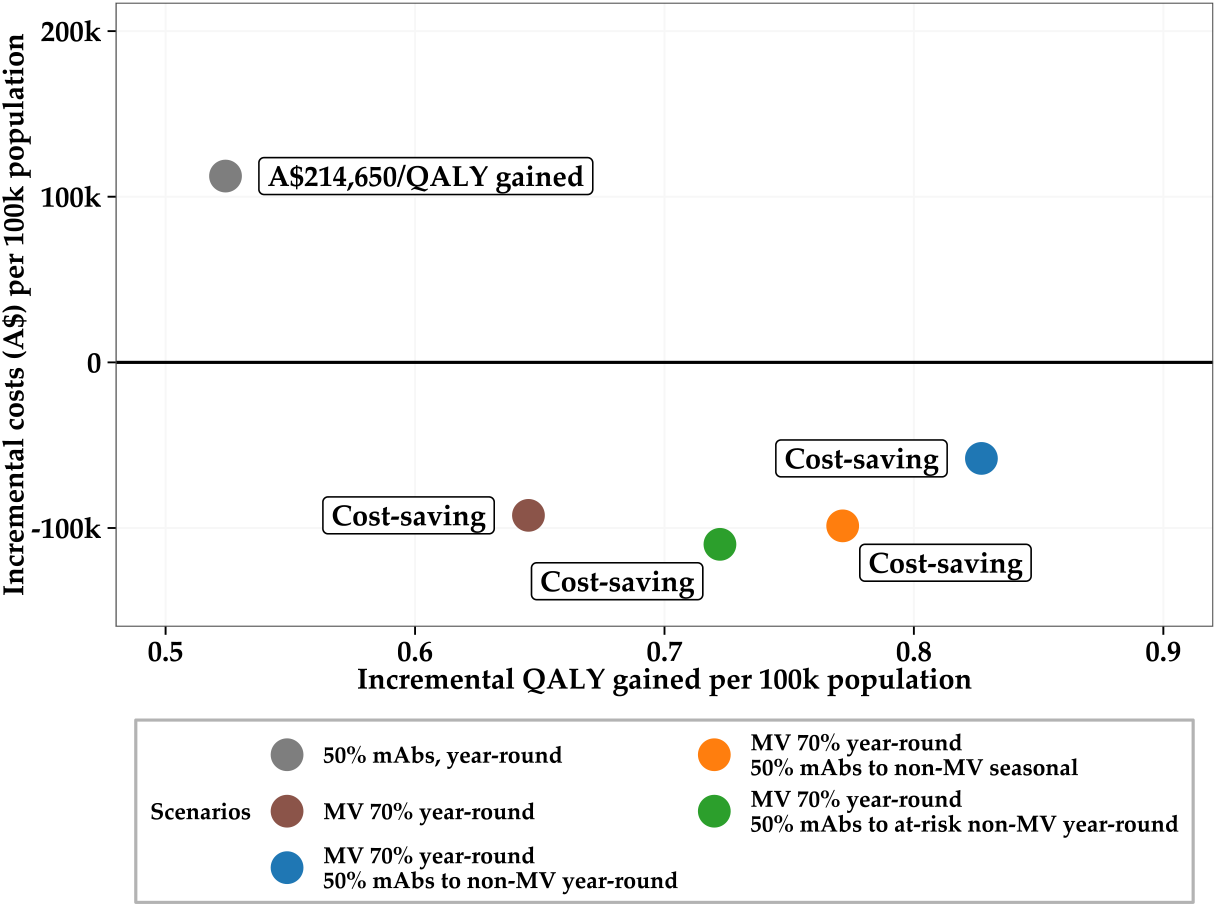
Mean incremental costs and incremental QALY gained per 100,000 population in scenarios using a scenario with only mAbs, a scenario with only MV and scenarios with combinations of mAbs and MV compared to no intervention. Abbreviations: mAbs = monoclonal antibodies; MV = maternal vaccination.

A one-way deterministic sensitivity analysis was conducted for the combination MV and mAbs scenario which delivered the greatest QALY gains (year-round MV delivered to 70% of pregnant people, with mAbs delivered year-round to 50% of newborns born without protection from MV). The ICER is most sensitive to the MV dose price, the cost per non-ICU hospitalisation and bias in the ascertainment of the hospitalisation burden (see Figure H.16 in the supplementary materials).

We varied the hospitalisation incidence and the assumed MV dose price together in a two-way sensitivity analysis (Figure 6), showing that if hospitalisation incidence is 50% higher than reported data suggest (i.e. 50% under-ascertainment in RSV-coded data, modelled via a multiplier of 1.5 on hospitalisation incidence), the program is cost-saving all the way up to a dose price of A$280 for the MV product. On the other hand, if hospitalisations are over-ascertained in reported data by 10% (a multiplier of 0.9), the MV dose price must be under A$130 for the program to be cost-saving.

**Figure 6.**
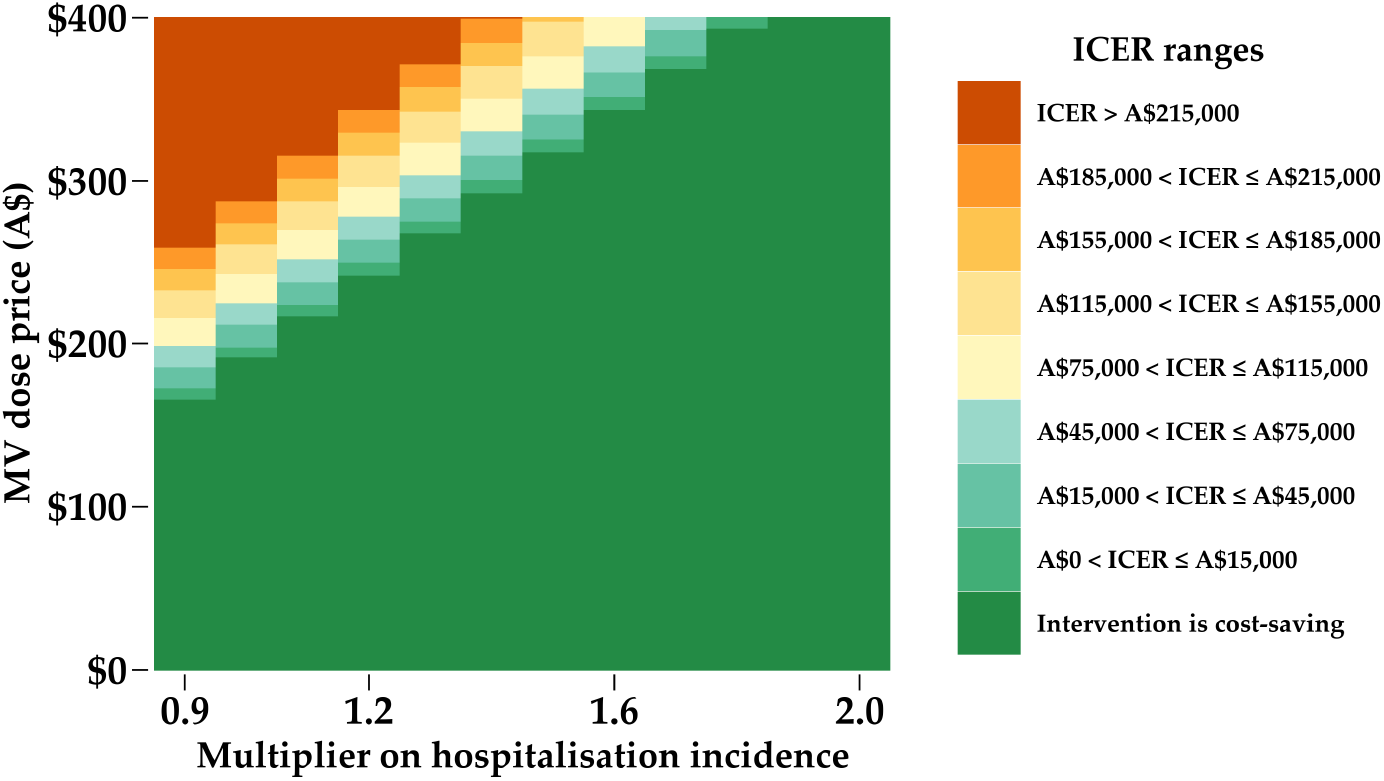
Two-way sensitivity analysis showing the impact on ICER (in terms of A$/QALY gained) of varying the assumed MV dose price and the hospitalisation incidence. The immunisation scenario is one in which 70% of pregnant people are given MV year-round, and 50% of newborns born without protection from MV are given mAbs year-round. Different colours correspond to different ranges of ICER. Abbreviations: MV = maternal vaccination; ICER = incremental cost-effectiveness ratio.

Panel A of Figure 7 presents the results of the Monte Carlo simulation-based probabilistic sensitivity analysis, illustrating the distribution of incremental costs and QALYs gained across a range of model simulations for the combined MV/mAbs vaccination scenario. The result suggests that this program is cost-saving in over 50% of simulations, however, there is considerable uncertainty as illustrated by the large spread of simulations. The range of potential incremental costs is largely driven by the range over which we sampled the MV dose price (see Appendix H in the supplementary materials). Panel B of Figure 7 builds on this analysis by quantifying the probability that the combined MV/mAbs scenario is cost-effective across a range of cost-effectiveness thresholds. Given the uncertainty in MV dose price and the large impact this has on cost-effectiveness, a sub-analysis was conducted in which MV dose price was kept fixed at three assumed values: $160 (as in our primary analyses), and the extrema of the plausible range. Results show that, despite uncertainty in all other model parameters, the combined vaccination scenario has a high probability of being cost-effective at MV dose prices of A$160 or less. However, at our highest estimate for the MV dose price (A$370), the program is much less likely to be cost-effective (<0.4 at a cost-effectiveness threshold of A$150k /QALY gained).

**Figure 7.**
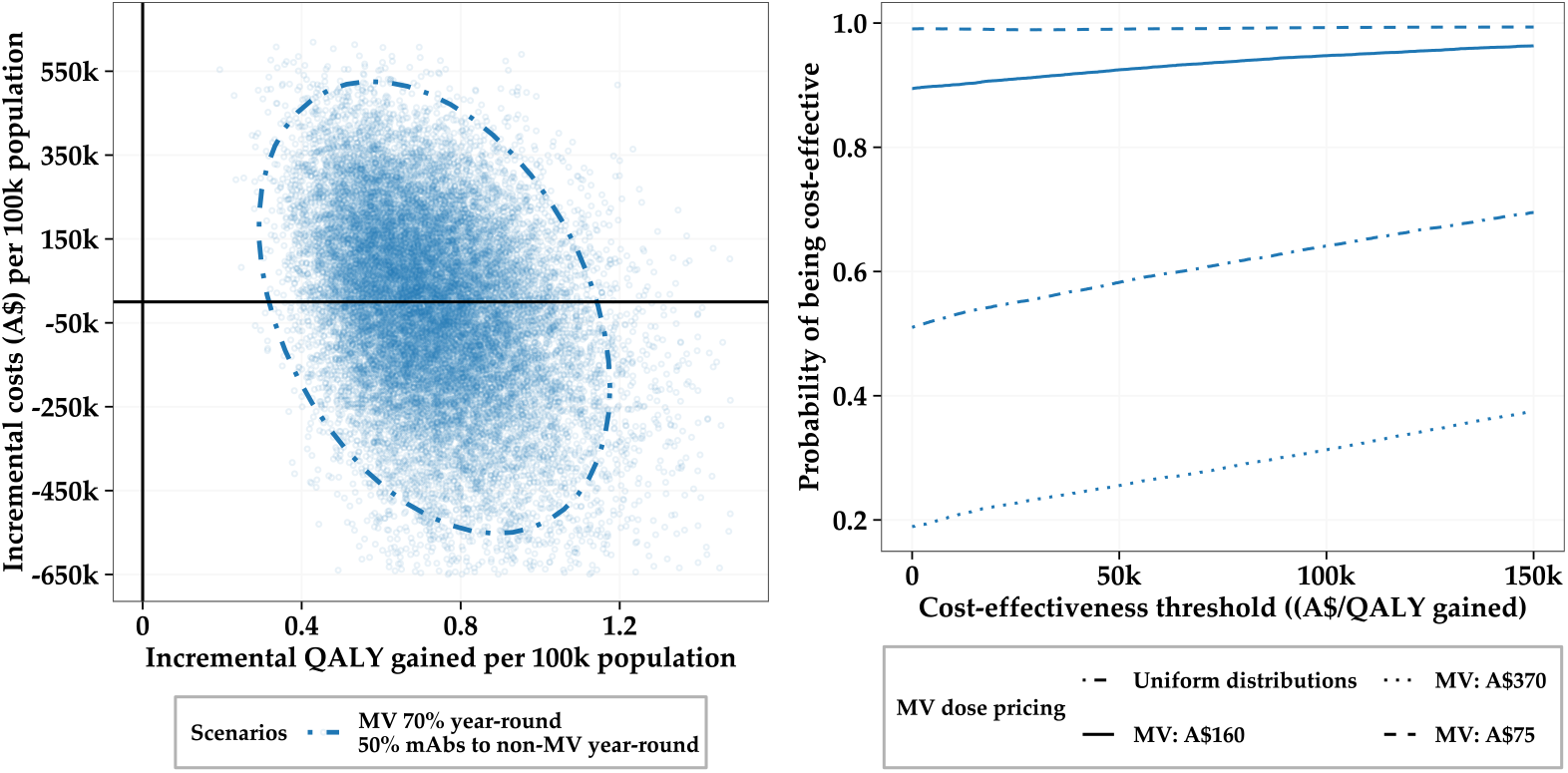
Results from the probabilistic sensitivity analysis for the scenario in which MV is delivered to 70% of pregnant people year-round, with mAbs delivered year-round to 50% of newborns born without protection from MV. Panel A: incremental costs and incremental QALYs gained per 100,000 population. Each dot represents a simulation result, with the dashed ellipse indicating the boundary encompassing 95% of simulations. Each simulation sampled parameter values from within the distributions outlined in Table H.12 in the supplementary materials. Panel B: Probability of the scenario being cost-effective across a range of cost-effectiveness thresholds. The MV dose price was either varied between A$75 and A$370 with a uniform distribution (dot-dash line) or kept fixed at A$75 (dashed line), A$160 (solid line), or A$370 (dotted line), while all other parameters were sampled from the distributions outlined in Table H.12. Abbreviations: QALY = quality-adjusted life years; MV = maternal vaccination; mAbs = monoclonal antibodies.

## 4. Discussion

We developed a pipeline of models to evaluate the likely impact of immunisation programs using MV and mAbs on RSV disease in infants over a two-year time horizon compared to no intervention. We showed that feasible programs combining both products (MV and mAbs) would substantially reduce the burden of RSV disease in Australian infants, and are likely to be cost-saving overall, from a healthcare system perspective. Our model was a key source of information, developed in close consultation with ATAGI and the Australian Government, to support national policy decisions on RSV immunisation. As of 2025, these programs are currently being implemented as outlined in the Australian Immunisation Handbook [28]. The RSV prevention program for infants in Australia is funded by separate processes; maternal vaccines are funded via the Commonwealth through the NIP, and long-lasting monoclonal antibodies are funded via individual states and territories [28].

Our results should be interpreted in the context of the uncertainty around our data inputs, particularly those relating to the healthcare burden of RSV disease in infants in our setting. In Australia, RSV only became a nationally notifiable disease in 2021. As such, there are limited high-quality national data on the annual number of medically-attended cases in each of our modelled healthcare settings, and in the transition probabilities between them.

In Australia, hospitalisation data are routinely coded to support analyses of burden of disease and collated by the AIHW. RSV-coded hospitalisations, as a proxy for severe RSV disease are likely well-ascertained in the health system — hospitalisations in infants have remained relatively stable from 2006 through 2015 [3], although we note a slight increase (approximately 20%) in coded hospitalisations in 2018–2019 data compared to earlier years. Notably, these data may include positive test results for RSV, in addition to hospitalisations coded as RSV based on clinical judgement. Thus, these data may not accurately represent the burden of disease. This uncertainty lead us to consider the possibility of both under- and over-ascertainment through a multiplier on the burden in our sensitivity analyses.

Further to changes to surveillance practices in recent years described above, quantifying the current burden is challenging given COVID-19 impacts. Public health and social measures implemented for COVID-19 led to changes to, and increases in, respiratory virus testing accessibility and practice, and disrupted RSV circulation in 2020–2022. Considering recent changes to respiratory virus surveillance in Australia, ongoing collection and analysis of age-specific, granular information on RSV infection and disease in all medically-attended clinical endpoints is critical for future modelling studies, and to assess the real-world effectiveness of RSV immunisation programs. This is important as the cost-effectiveness for our combination scenario (as currently recommended and being implemented by the Australian Government) is particularly sensitive to multiple sources of uncertainty, including potential over- or under-ascertainment of hospitalisations, and the assumed protective efficacy of immunising products against clinical endpoints.

In addition to the baseline incidence of RSV cases and hospitalisations, uncertainty around the MV dose price was a large driver of the cost-effectiveness of the combined MV/mAbs immunisation scenario. Increasing the dose price of MV from A$180 to A$250 increases the ICER from cost-saving to over A$190,000/QALY gained, if we have under-ascertained the hospitalisation burden by 10%. The dose price at which the program is cost-saving depends strongly on the assumed baseline burden of disease, highlighting the importance of continued surveillance to support evidence-based policy-setting.

The body of international evidence exploring the cost-effectiveness of new maternal and infant-immunising products for RSV is growing rapidly. The majority of published studies use static cohort models and relatively few evaluate combined maternal and infant immunisation strategies [29, 30]. A strength of our work is the use of an individual-based dynamic transmission model, which allows tracking of risk status, infection histories, and delivery of interventions at the individual level. Notably, linking pregnant-person–infant pairs enables modelling of feasible combinations of immunisation programs that selectively target infants who are not protected through maternal vaccination programs. Studies in other high-income countries (the United Kingdom and Canada) of the impact and cost-effectiveness of combination strategies have also found that combination strategies are likely cost-effective, if not cost-saving [7, 31, 23]. Generally, these studies found that the scenarios most likely to be cost-effective were those that involved seasonal administration for either product, followed by seasonal catch-up for mAbs which are best-targeted to at-risk infants. One study in Canada found that combination programs of mAbs and MV were not cost-effective. However, similar to our findings, dose price was a major driver of these results; such that if their MV dose price was reduced to CAD $60–125 (A$65–140, noting our base-case MV dose price was A$160) some combination scenarios became cost-effective [32].

Seasonal MV programs were not considered in our modelling, due to logistical and equity considerations, aligning with MV programs for other infectious diseases in Australia. In any case, our results show that year-round MV programs are likely cost-effective in Australia, concordant with another cost-effectiveness study from Australia which used a dynamic transmission model [8].

We did not ascribe significant protective efficacy against infection for either immunising product for infants. There are limited data from clinical trials or emerging real-world studies to distinguish efficacy against medically-attended endpoints and efficacy against infection. We made the conservative choice to apply the bulk of the protective effect of the immunising products to preventing severe clinical endpoints. This meant we did not see any indirect effects in infants who did not receive an immunising product. In the absence of supporting evidence, other modelling studies have assumed that these products confer a higher protection against infection, and have consequently identified potential indirect effects, which reduced hospitalisations in infants by 22–30% [11, 33]. We also do not model any direct effects in the pregnant people who receive MV, given a similar lack of evidence. As the RSV burden is considerably lower for people of child-bearing ages compared to infants, we do not expect that implementing any direct effects would significantly alter our assessment of the cost-effectiveness of programs involving MV. As modelling direct effects in adults receiving vaccination would result in a larger averted RSV disease burden, scenarios involving MV would become even more cost-saving.

An additional limitation of our modelling analysis is that we excluded potential long-term sequelae of RSV infection in infancy. RSV infection in infants is associated with increasing the risk of developing asthma [34, 35], and thus, we may be underestimating the impact and cost-effectiveness of RSV-immunising products. Future analyses could incorporate emerging data on the impact of these immunisation products on reducing the risk and burden of asthma later in life, to provide a more comprehensive assessment.

## 5. Conclusions

This modelling and cost-effectiveness analysis has shown that combination immunisation programs using mAbs and MV are likely to reduce the burden of RSV disease in Australian infants and be cost-saving from a population perspective. While the results indicate that combination programs are likely cost-saving, we note that the results depend strongly on the uncertain dose price for both mAbs and MV, e.g. increasing the dose price of MV from A$160 to A$250 increases the ICER from cost-saving to over A$200,000/QALY gained. Our analysis has informed ATAGI and Australian Government recommendations for an RSV immunisation program to protect newborns and infants.

## Data Availability

All data produced in the present work are contained in the manuscript or are available online in the code repository at https://gitlab.unimelb.edu.au/julian.carlin/infant-rsv-modelling.

## 6. Acknowledgements

We thank the Australian Technical Advisory Group on Immunisation (ATAGI) secretariat and members, especially those within the RSV/respiratory working groups (chaired by A/Prof. Katherine Gibney) for provision of technical advice to support the development of this model. The ATAGI member list as of 2025 is available at https://www.health.gov.au/committees-and-groups/atagi/members.

We would like to particularly acknowledge Jocelynne McRae, Jean Li-Kim-Moy, Sanjay Jayasinghe and Bette Liu from the National Centre for Immunisation Research and Surveillance for working closely with us to provide access to AIHW-supplied data and in-confidence analyses. Similarly, we acknowledge Jane Tuckerman, Danielle Wurzel and Nigel Crawford from the Murdoch Children’s Research Institute for provision of paediatric data from the Royal Children’s Hospital, Melbourne.

## 7. Declaration of interest

Authors declare no conflicts of interest.

## Notes

### Competing Interest Statement

The authors have declared no competing interest.

### Author Declarations

This study was funded by the National Immunisation Division of the Australian Government Department of Health, Disability and Ageing to support policy decision making. The model was parameterised using nationally aggregated patient data provided by the Department of Health, Disability and Ageing using Admitted Patient Care data supplied by the Australian Institute of Health and Welfare (AIHW). National Admitted Patient Care data (i.e. line by line unit records), collated by AIHW and supplied to the Department of Health, Disability and Ageing, were aggregated and suppressed by the Department before being provided to the authors. Approval for the use of these data for analyses and subsequent publication was granted by the Department. Clinical risk factors in early infancy were parameterised using de-identified patient data from the Royal Children's Hospital, use of which was granted by the Murdoch Children's Research Institute Ethics Committee (#37185)

