## Supplemental Materials for "Cost-effectiveness of immunising interventions to reduce respiratory syncytial virus disease burden in infants in Australia"

### Appendix A. Demography

The transmission model consists of $10^5$ individuals with ages and sexes initially distributed according to Australian Bureau of Statistics (ABS) data from 2012 [1]. As these data only contain information for individuals up to 99 years-of-age we set that as the maximum age in our model. Deaths occur probabilistically in the model, with age- and sex-specific rates listed in ABS life table data from 2012–2014 [2]. We initialise the population using data from 2012, as we "burn in" the model for roughly five and a half years to initialise the history of prior infection appropriately and stabilise disease dynamics. The age-sex distribution input into the model is visualised in the left panel of Figure A.1. The age-at-death distribution is visualised in the right panel of Figure A.1, where only deaths that occur in the next five modelled years are shown. We do not include migration in the model, as over the short time horizon on which the model is run this factor is not relevant.

We model pregnancy and birth in the model. We assume that 89%[3] of women between the ages of 15 and 49 (inclusive) are eligible to become pregnant. We then apply age-specific fertility rates, reported in ABS data from 2022 [4] to choose which eligible people become pregnant in the model. Gestational ages are selected randomly from a distribution derived from Australian Institute of Health and Welfare (AIHW) 2021 reporting [5]. The age-specific fertility rates are shown in the left panel of Figure A.2. The distribution of gestational ages is shown in the right panel of Figure A.2. For simplicity, we do not model multiple-births or stillbirths. When modelling scenarios that include vaccines delivered to pregnant people these

[1]Co-first author
[2]Co-senior author
[3]This value accounts for 11% of women who will not become pregnant in their lifetime, derived using ABS census data [3].

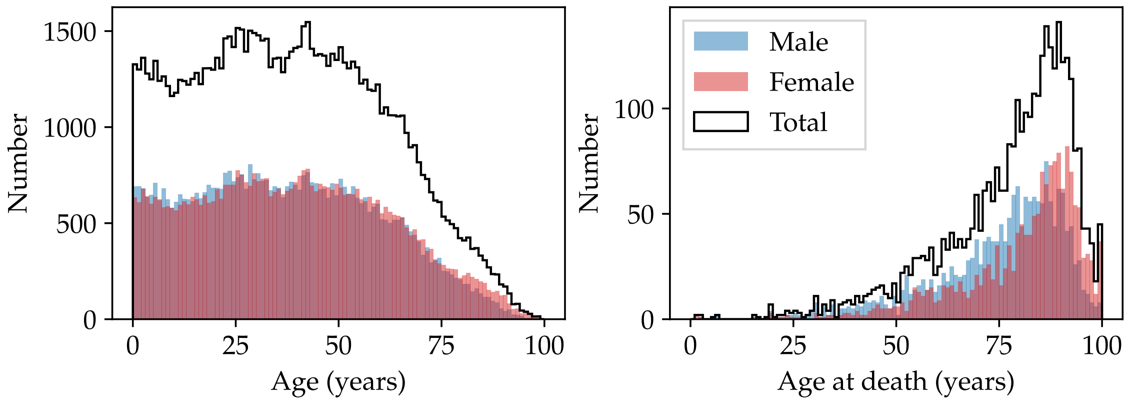


Figure A.1: Left panel: Number of males and females at different ages input into the whole-population model of size 100,000. Right panel: Number of deaths at different ages for the whole-population model, only considering deaths that occur in the next five modelled years.

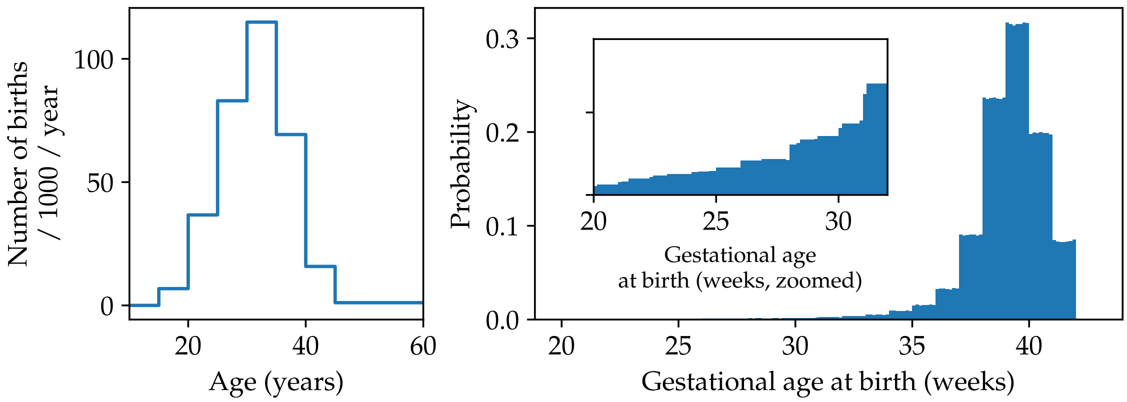


Figure A.2: Left panel: Age-specific fertility rates assumed in the model, sourced from ABS 2022 data. Right panel: Distribution of gestational ages, with inset panel emphasising that premature births are included in the modelling, sourced from AIHW 2021 reporting.

simplifications will impact the results slightly — multiple births would allow one maternal vaccine to benefit multiple newborns, increasing the effectiveness of a program with maternal vaccines, while on the other hand stillbirths that occur after the delivery of a maternal vaccine would lower the effectiveness of a program with maternal vaccines. The magnitude of these two effects is not equal, as the probability that a stillbirth occurs after 28 weeks of gestation is at most 0.4% [6]. The probability of multiple births is roughly 1.5% [4].

We also do not model household structure — the prevalence of community-spread RSV down-weights the importance of modelling within-household transmission [3].

### Appendix B. Transmission model details

#### *Appendix B.1. Compartments and contact matrix*

Each alive individual in the model is assigned one of four states: susceptible to infection (S), exposed and infected but not yet infectious (E), infectious (I), and temporarily immune while recovering (R). We visualise the four compartments in Figure B.3, noting the reduced susceptibility to infection following recovery (see

Appendix B.3 for details).

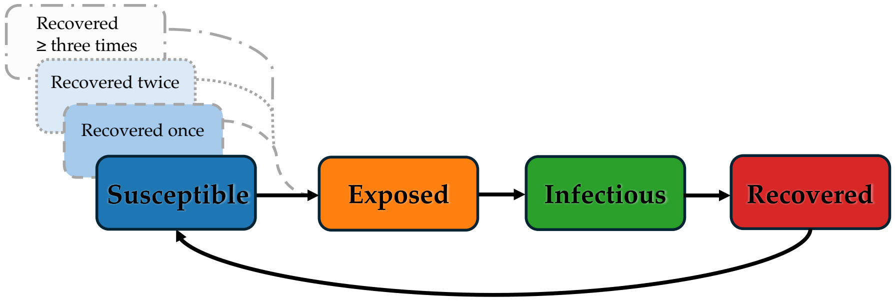


Figure B.3: Schematic illustrating the flow between disease compartments for each individual in the transmission model.

Not all individuals in the S state are equally likely to be infected. Given we do not explicitly model households (see Appendix A), contact between susceptible and infectious individuals is assumed to occur at a population-level. We use age-stratified mixing patterns as reported in Mistry et al. 2021 [7], which inform the average number of daily contacts that a person in Australia in a given one-year age group has with all other one-year age groups (up to 85 years of age). The contact matrix, $\boldsymbol{C}$, is shown in Figure B.4.

In the model, time is discretised into time-steps $t_i$, with $\Delta t \equiv t_{i+1} - t_i$. Unless otherwise noted we use $\Delta t = 1$ day. Every time-step, the number of individuals in I is computed for each age-group in the contact matrix, producing a row-vector $\boldsymbol{N}_{\mathrm{I}}(t_i)$. As $\boldsymbol{C}$ has units of contacts per day, the number of possible transmission events per age-group in the time-step $\boldsymbol{N}_{\mathrm{PT}}(t_i)$ is

$$\boldsymbol{N}_{\mathrm{PT}}(t_i) = \frac{1}{\Delta t} \boldsymbol{C} \cdot \boldsymbol{N}_{\mathrm{I}}(t_i) \,. \tag{B.1}$$

These possible transmission events are then assigned randomly to individuals within each age-group. If the assigned individuals are in the S state, they are then probabilistically assigned to the E state. The probability of transitioning from S to E is governed by four factors:

i) the probability of infection given contact, $\beta(t_i)$, described further in Appendix B.2;

ii) the protection against acquisition due to antibodies built up from prior infections, described further in Appendix B.3;

iii) the protection against acquisition due to short-lived maternal antibodies from cross-placental transfer, described further in Appendix B.4; and

iv) the protection against acquisition due to mAbs or MV, as described in Appendix B.5.

Each individual that transitions from S to E will spend a random amount of time in the E state before transitioning to the I state. The random time is drawn from a Gamma distribution with a mean of 4 days and a shape parameter of 3. The length of time spent in the I state is also random, following a Gamma distribution with shape parameter 3, but with a mean of 9 days. These distributions are guided by literature sources [8–10]. After becoming no longer infectious the individual moves into the R state. The length of

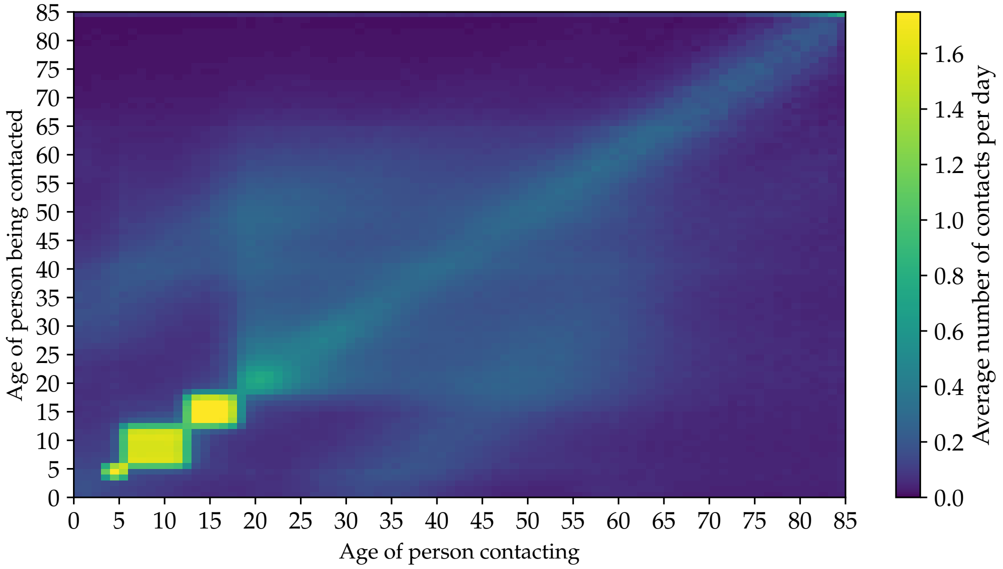


Figure B.4: Average number of contacts per day used in the transmission model. Data from Mistry et al. 2021 [7].

time completely immune (i.e. before transitioning back to the S state) is also random, and is drawn from a Gamma distribution with shape parameter 3, and a mean of $\mu_{\mathrm{R}}$ days. The value of $\mu_{\mathrm{R}}$ is determined via model calibration (see Appendix C for details).

*Appendix B.2. Seasonality*

The seasonal nature of RSV epidemics is encoded in the model through two choices: the period of time for which individuals are completely immune after recovering from a previous infection, and the seasonal forcing term in the probability of infection given contact, viz.

$$\beta(t_i) = \beta_0 \left[ 1 + \beta_1 \sin\left( \frac{2\pi\, t_i}{365 \text{ days}} \right) \right] , \tag{B.2}$$

where $\beta_0$ is the average probability of infection given contact, $\beta_1$ is the magnitude of the seasonality adjustment. We adjust the phase of the seasonality by "burning in" the transmission model for $t_{\mathrm{burn}}$ days. The values of $\beta_0$, $\beta_1$, and $t_{\mathrm{burn}}$ are determined via model calibration (see Appendix C for details).

*Appendix B.3. Protection against acquisition of infection due to repeated recoveries*

To capture age-specific incidence of infection, we assume that after the length of time spent completely immune in R, an individual moves from R to S, where they have partial protection against infection, dependent on the number of previous recoveries. This partial protection is encoded as a probability, $p_{\mathrm{prot.}}$, for a potential transmission event to *not* result in an infection. This protection wanes with a half-life $\tau_{\mathrm{prot.}}$, viz.

$$p_{\mathrm{prot.}}(t, t_{\mathrm{recov.}}, j) = p_{j,\,\mathrm{min}} + (p_{j,\,\mathrm{max}} - p_{j,\,\mathrm{min}}) \exp\left[ -\log 2 \left( \frac{t - t_{\mathrm{recov.}}}{\tau_{\mathrm{prot.}}} \right) \right] , \tag{B.3}$$

where $t_{\mathrm{recov.}}$ is the time that the individual moved from R to S, $j$ is the number of previous recoveries they have experienced, $p_{j,\,\mathrm{max}}$ is the maximum protection just after the $j$-th recovery, and $p_{j,\,\mathrm{min}}$ is the minimum

Table B.1: Parameters assumed for the build-up of probabilistic protection against subsequent acquisition of infection. On transition from R to S, protection is set at $p_{j,\,\mathrm{max}}$, based on the number of previous recoveries $j$, and begins to wane thereafter. Long-term protection is the asymptotic value as the number of days since last recovery gets large ($\gg \tau_{\mathrm{prot.}}$).

| Number of previous recoveries | Initial protection $p_{j,\,\mathrm{max}}$ | Long-term protection $p_{j,\,\mathrm{min}}$ |
|---|---|---|
| 1 | 0.5 | 0 |
| 2 | 0.75 | 0.25 |
| 3+ | 1 | 0.5 |

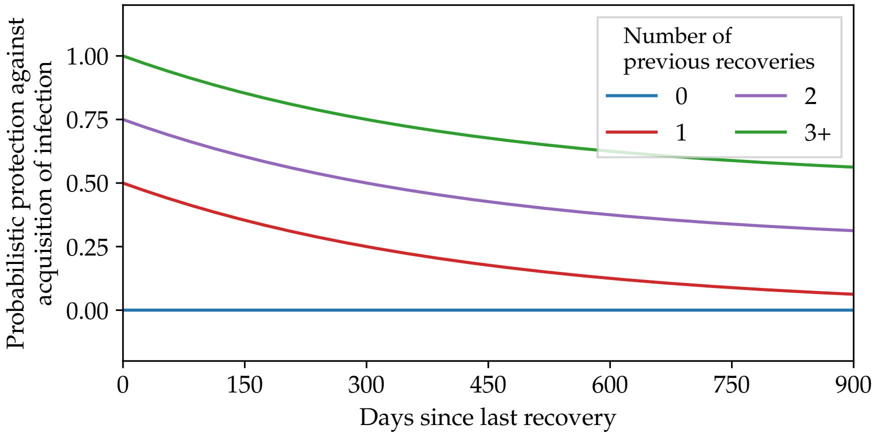


Figure B.5: Assumed protection against acquisition of infection due to prior infections.

protection a long time ($t - t_{\mathrm{recov.}} \gg \tau_{\mathrm{prot.}}$) after the $j$-th recovery. We tabulate the assumed values of $p_{j,\,\mathrm{min}}$ and $p_{j,\,\mathrm{max}}$ in Table B.1. We assume $\tau_{\mathrm{prot.}} = 300$ days, guided by Ref. [11]. We visualise this build-up of protection from repeated infection and recovery in Figure B.5. We find that 95% of individuals have experienced three recoveries by 5.5 years-of-age.

We emphasise that while it captures some facets of antibody build-up from repeated infection, the protection against infection modelled in Equation (B.3) does not impact the clinical pathways model described in Section 2.2. That is, it does not alter the severity of an RSV infection, only the probability of acquisition of infection.

*Appendix B.4. Protection against acquisition of infection in newborns due to maternal antibodies from prior infections*

Due to the observed phenomenon of cross-placental transfer of maternal antibodies, we assign a short-lived protection against acquisition of infection to newborns, $p_{\mathrm{newborn}}$, scaled to their pregnant person's partial protection at time of birth $p_{\mathrm{prot.}}(t_{\mathrm{birth}}, t_{\mathrm{recov.}}, j)$, see Equation (B.3). Explicitly, we assign $p_{\mathrm{newborn},\,0} = p_{\mathrm{prot.}}(t_{\mathrm{birth}}, t_{\mathrm{recov.}}, j)$, and assume this partial protection wanes to zero with a half-life of $\tau_{\mathrm{newborn\ prot.}}$ viz.

$$p_{\mathrm{newborn}}(t, t_{\mathrm{birth}}) = p_{\mathrm{newborn},\,0} \exp\left[-\log 2 \left(\frac{t - t_{\mathrm{birth}}}{\tau_{\mathrm{newborn\ prot.}}}\right)\right] . \tag{B.4}$$

We assume $\tau_{\mathrm{newborn\ prot.}} = 35$ days, guided by Refs. [12–15]. If an infant is infected and recovers, the protection against subsequent infection is calculated via Equation (B.3) instead of Equation (B.4).

As for $p_{\text{prot.}}$, we reiterate that while the above protection against acquisition of infection acts in a similar way to antibodies in preventing infection, it does not impact the clinical pathways model described in Section 2.2. We explicitly separate out the protection against acquisition of infection and protection against severe disease outcomes as we do not have adequate evidence to map antibody levels to severity of disease.

*Appendix B.5. Protection against acquisition of infection due to interventions*

Clinical trials for MV and mAbs do not directly measure protection against acquisition of infection [16–27]. In the absence of other data, for the infant products we use a fixed percentage of the efficacy that is applied against GP visits (see Appendix D.5) as the efficacy against acquisition of infection. Explicitly, we assume an initial efficacy $\varepsilon_{\text{mAbs}}$ of 10% against acquisition of infection for mAbs, and an initial efficacy $\varepsilon_{\text{MV}}$ of 6% against acquisition of infection for MV. The data from clinical trials also does not clearly lay out the duration of protection afforded by the products. We assume that mAbs have constant $\varepsilon_{\text{mAbs}}$ for $\tau_{\text{mAbs, const.}} = 90$ days after the product is delivered to the infant at time $t = t_{\text{dose}}$, then begin to wane with a half-life of $\tau_{\text{mAbs, h.l.}} = 70$ days [17, 15], viz.

$$\varepsilon_{\text{mAbs}}(t) = \begin{cases} \varepsilon_{\text{mAbs}}(0) & \text{, if } t - t_{\text{dose}} < \tau_{\text{mAbs, const.}} \\ \varepsilon_{\text{mAbs}}(0)\, \exp\left[-\log 2 \left(\dfrac{t - t_{\text{dose}}}{\tau_{\text{mAbs, h.l.}}}\right)\right] & \text{, otherwise.} \end{cases} \tag{B.5}$$

We assume that MV have a constant $\varepsilon_{\text{MV}}$ for $\tau_{\text{MV, const.}} = 90$ days after the product is delivered to the pregnant person at time $t = t_{\text{dose}}$, then begin to wane linearly over the subsequent $\tau_{\text{MV, lin.}} = 180$ days [23]., viz.

$$\varepsilon_{\text{MV}}(t) = \begin{cases} \varepsilon_{\text{MV}}(0) & \text{, if } t - t_{\text{dose}} < \tau_{\text{MV, const.}} \\ \varepsilon_{\text{MV}}(0)\, \dfrac{\tau_{\text{MV, lin.}} - (t - t_{\text{dose}} - \tau_{\text{MV, const.}})}{\tau_{\text{MV, lin.}}} & \text{, if } 0 \le t - t_{\text{dose}} - \tau_{\text{MV, const.}} < \tau_{\text{MV, lin.}} \\ 0 & \text{, otherwise.} \end{cases} \tag{B.6}$$

The efficacy against acquisition of infection described in Equations (B.5) and (B.6) both provide a (small) probability of avoiding infection, following the protection applied via Equation (B.3). That is, these protections are effectively multiplicative. This interaction is visualised for an exemplar newborn in Figure B.6. We apply the efficacy of MV and mAbs in this manner as the protection afforded by Equation (B.3) is the underlying "base case" for the population, i.e. clinical trials measure efficacy in a population of people who have naturally been infected in the past. The above parameters and modelling choices were made via an expert elicitation process with experts in the Australian Technical Advisory Group on Immunisation (ATAGI) respiratory virus working group.

The efficacy described in this section only pertains to protection against acquisition of infection. We discuss the efficacy against clinical endpoints in Appendix D.5.

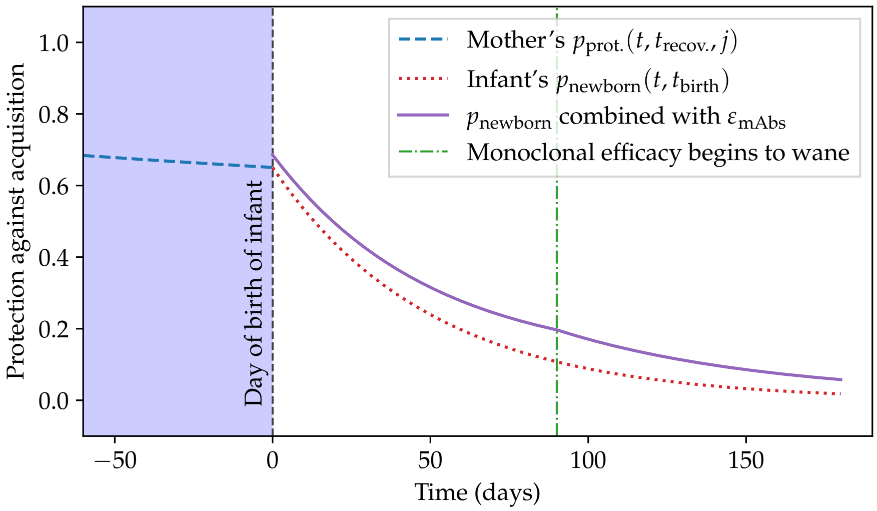


Figure B.6: Visualising the modelling choices described in Appendix B.3 and Appendix B.5 for an exemplar newborn who receives a dose of mAbs at birth.

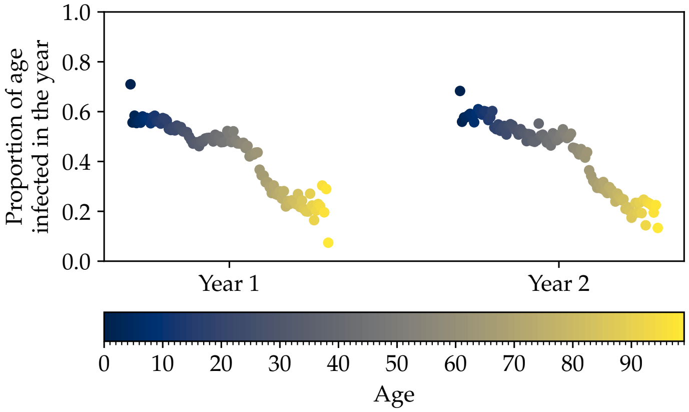


Figure C.7: Average proportion of each age infected in two representative years of simulated RSV transmission.

### Appendix C. Calibration

Model calibration aims to match two main elements of RSV infection and transmission in Australia. These are:

i) The average incidence of infection for children should match the accepted heuristic that roughly 75% of infants are infected with RSV by one year of age, and roughly 95% by two years of age [28, 29]; and

ii) infections should have regular yearly epidemic waves across all age groups, with the magnitude and duration of the waves determined by the monthly hospitalisation records, as described in more detail in Appendix D.

There are four main epidemiological parameters in the model that are unguided by the literature, namely $\beta_0$, $\beta_1$, $t_{\text{burn}}$, and $\mu_{\text{R}}$, see Appendix B for definitions of these parameters. Alongside these parameters we simultaneously determine the functional form of the mapping between infection and hospitalisation, i.e. $p(\text{H}^+)$, so that we can compare modelled infections with observed hospitalisation data. For the latter, we are guided by the observation that infants and the elderly have an increased incidence rate ratio (IRR) compared to other ages, suggesting a "bath-tub" shape for $p(\mathrm{H}^+)$. For children up to two years of age, we model this curve as a sigmoid, introducing four additional parameters $\boldsymbol{\theta}$, viz.

$$p(\mathrm{H}^+ \mid \mathrm{age}, \boldsymbol{\theta}) = p_{\min} + (p_{\max} - p_{\min}) \left\{ 1 + \frac{\exp(-ab)}{1 + \exp\left[-a\left(\mathrm{age} + b\right)\right]} \right\} \text{ if age} < 2\,\text{years}, \tag{C.1}$$

where $\boldsymbol{\theta} = \{p_{\min}, p_{\max}, a, b\}$.

The resultant best-fit parameters from a human-supervised, iterative model-fitting procedure are shown in Tables C.2 and C.3. We show the average proportion of each age infected per year in Figure C.7. We compare the modelled monthly average hospitalisations by age group and the observed monthly hospitalisation data in Figures 3, in the main text. The resultant shape of Equation (C.1) with parameters listed in Table C.3 is shown as the red curves in Figures D.8.

Table C.2: Epidemiological parameters determined during the calibration process.

| Parameter | Description | Value |
|---|---|---|
| $\beta_0$ | Base probability of transmission given contact, see Equation (B.2) | 0.06 |
| $\beta_1$ | Seasonal variation in probability of transmission given contact, see Equation (B.2) | 0.01 |
| $t_{\mathrm{burn}}$ | Burn-in duration to correctly offset seasonality | 2044 days |
| $\mu_{\mathrm{R}}$ | Mean duration of complete immunity following an RSV infection | 310 days |

### Appendix D. Clinical pathways model details

#### *Appendix D.1. Data and conditional probabilities*

For the less-severe outcomes: not seeking medical care (nSMC), GP, and ED, there are limited Australian data to inform the appropriate probabilities of ending at these outcomes, given infection. For the more-severe outcomes: hospitalisation (H), ICU, and death (F) we have used Australian data. National Admitted Patient Care data (i.e. line by line unit records), collated by AIHW and supplied to the Department of Health, Disability and Ageing, were aggregated and suppressed by the Department before being provided to the authors.

Table C.3: Parameters determined during the calibration process for the age-specific $p(\mathrm{H}^+)$ in Equation (C.1).

| Parameter | Value |
|---|---|
| $p_{\max}$ | 0.08 |
| $p_{\min}$ | 0.01 |
| $a$ | −1.75 |
| $b$ | −0.5 |

In the mathematical description below, a superscript of $^+$ denotes "at least this clinical endpoint". Almost all probabilities considered are age- and risk-status-specific; the explicit dependence on these properties is suppressed for now.

The aggregated data contains, by month-of-age and month of the year:

i) the number of individuals who are admitted to hospital — using International Statistical Classification of Diseases and Related Health Problems, Tenth Revision, Australian Modification (ICD10AM) codes for principal diagnoses (J21.9) and principal or additional diagnoses (J12.1, J20.5, B97.4, J21.0), $N_{\mathrm{H}^+}$, per Saravanos et al. 2019 [30]

ii) the number of individuals who are admitted to ICU — ICD-10-AM diagnosis codes per above, $N_{\mathrm{ICU}^+}$,

iii) the number of individuals who die, after being admitted to ICU (or from hospitalisation) — ICD-10-AM diagnosis codes per above, $N_{\mathrm{F}}$.

We use i) as part of model calibration. We describe the calibration process in detail in Appendix C. In summary, we iteratively adjust epidemiological parameters, and the parameters that describe the shape of the infection-hospitalisation rate as a function of age. This iteration continues until we can match modelled hospitalisations to observed hospitalisations. For the purposes of the clinical pathways model, this iterative calibration process produces a parameterised description of the probability of at-least-hospitalisation given infection $p(\mathrm{H}^+)$, as a function of age.

We assume that all infections for which ICU is the final clinical endpoint will also be counted in the number of hospitalisations, i.e. all individuals counted in $N_{\mathrm{ICU}^+}$ are also counted in $N_{\mathrm{H}^+}$. That is, we estimate the probability of ICU admission (or worse) given hospitalisation, viz.

$$p(\mathrm{ICU}^+ \mid \mathrm{H}^+) \approx \frac{N_{\mathrm{ICU}^+}}{N_{\mathrm{H}^+}} \,. \tag{D.1}$$

We also assume that all infections for which F is the final clinical endpoint will also be counted in the number of ICU admissions. The probability of fatality, given ICU admission is estimated as

$$p(\mathrm{F} \mid \mathrm{ICU}^+) \approx \frac{N_{\mathrm{F}}}{N_{\mathrm{ICU}^+}} \,. \tag{D.2}$$

We find that $p(\mathrm{F} \mid \mathrm{ICU}^+) \approx 0.0036$ (95% CI: 0.0012–0.0084, 5/1385). We do not model an age-specific $p(\mathrm{F} \mid \mathrm{ICU}^+)$ due to the small number of fatalities in granular month-of-life age groups.

The less-severe clinical outcomes of nSMC, GP, and ED, are less well-informed by Australian data. Takashima et al. (2021) [31], which followed a birth-cohort in Queensland for two years (2010–2012) with weekly sampling irrespective of symptoms, found that 34% (95% CI: 24%–45%, 28/82) of infants sought medical care (SMC) given RSV infection. Modelling from the UK provides additional data points at five other age points, up to five years of age [32][4]. The same summary of UK modelling also helps inform the age-specific probability

[4] See Table 7 in the Supplementary Materials 1 of Ref. [32].

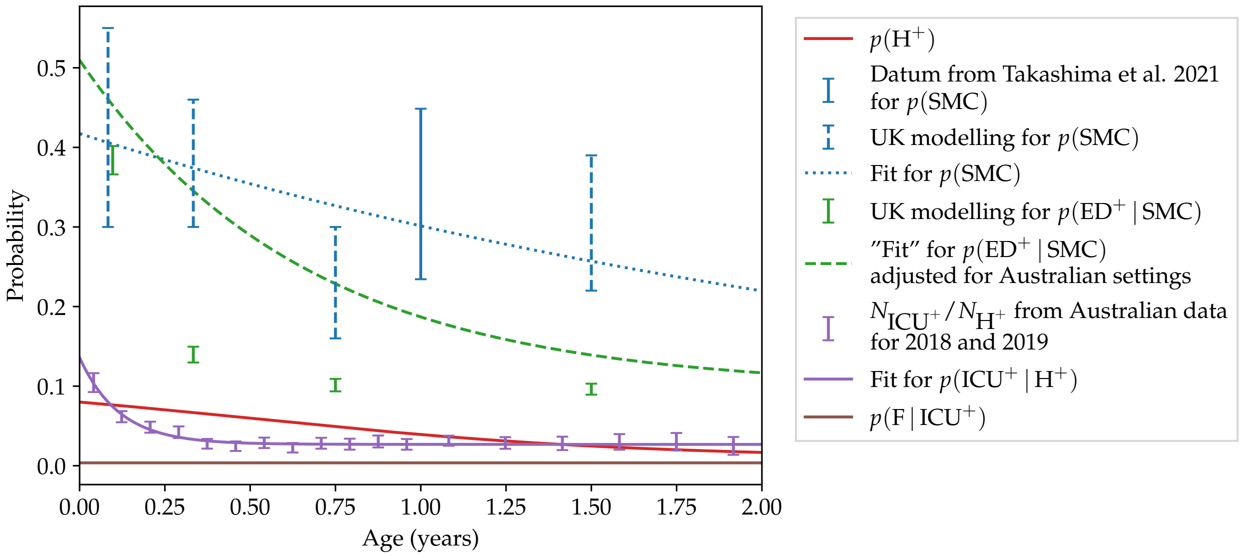


Figure D.8: Conditional transition probabilities for the infant clinical pathways model, and associated data with which they are informed.

of ED presentation given that medical care is sought, $p(\mathrm{ED} \mid \mathrm{SMC})$. However, to account for differences in settings of care between the UK and Australia, we assume values of $p(\mathrm{ED}^+ \mid \mathrm{SMC})$ to be larger at all ages up to two years-of-age. That is, we assume that proportionally more infants seek care at ED over the GP in Australia, as compared to infants in the UK. This was determined in consultation with clinical experts on ATAGI.

To capture the age-specific nature of $p(\mathrm{ICU}^+ \mid \mathrm{H}^+)$, $p(\mathrm{ED}^+ \mid \mathrm{SMC})$ and $p(\mathrm{SMC})$ we fit an exponential through the available data for each age-specific conditional probability for infants aged up to two years-of-life. Details are in Appendix D.2. The form of $p(\mathrm{H}^+)$ as a function of age is detailed in Appendix C.

For the purposes of the clinical pathways model we convert these conditional, age-specific probabilities to the absolute probabilities of ending at each endpoint. The algebra is expanded in Appendix D.3. We show the conditional transition probabilities in Figure D.8 and the absolute probabilities in Figure D.9.

*Appendix D.2. Smoothing conditional probabilities*

As discussed in Appendix D.1, we fit smooth curves to data that inform various conditional probabilities as a function of age (up to two years-of-age).

All of $p(\mathrm{SMC})$, $p(\mathrm{ED}^+ \mid \mathrm{SMC})$, and $p(\mathrm{ICU}^+ \mid \mathrm{H})$ are well-fit with a decaying exponential of functional form

$$y = y_{\mathrm{min}} + \exp\left[ -\lambda \left( x - x_{\mathrm{offset}} \right) \right] . \tag{D.3}$$

The parameters $\{y_{\mathrm{min}}, \lambda, x_{\mathrm{offset}}\}$ are found via a non-linear least-squares optimisation algorithm implemented in `scipy.optimize.curve_fit`. The resultant parameters used in the clinical pathways model (see the blue-dotted, green-dashed, and purple-solid curves in Figure D.8) are tabulated in Table D.4.

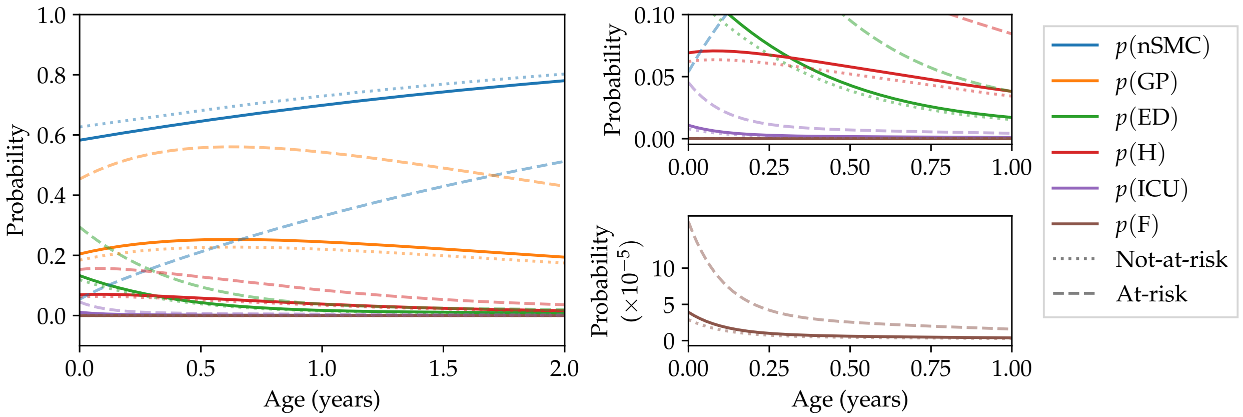


Figure D.9: Probability of different clinical endpoints as a function of age (up to two years-of-life). Panels on the right show zoomed regions of the main panel, to emphasise that all of these probabilities are age-specific. The probabilities for not-at-risk infants are shown as dotted curves, while the probabilities for at-risk infants are shown as dashed curves, see Section Appendix D.4 for details.

Table D.4: Parameters used to model the smooth conditional probabilities in the clinical pathways model that can be represented with an exponential, see Equation (D.3) for the functional form.

| Infants | $y_{\min}$ (probability) | $\lambda$ (years$^{-1}$) | $x_{\text{offset}}$ (years) |
|---|---|---|---|
| $p(\text{SMC})$ | 0.025[1] | 0.350 | −2.67 |
| $p(\text{ED}^+ \mid \text{SMC})$ | 0.0972 | 1.53 | −0.581 |
| $p(\text{ICU}^+ \mid \text{H}^+)$ | 0.0268 | 8.58 | −0.258 |

[1] This parameter is fixed *a priori* to ensure the probability of SMC does not become negative at higher ages.

#### *Appendix D.3. Converting conditional to absolute endpoint probabilities*

As described in Appendix D.1 we have data to inform the following conditional transition probabilities: i) $p(\text{SMC})$, ii) $p(\text{ED}^+ \mid \text{SMC})$, iii) $p(\text{H}^+)$, iv) $p(\text{ICU}^+ \mid \text{H}^+)$, and v) $p(\text{F} \mid \text{ICU}^+)$.

For simplicity in sampling the clinical pathways model (and to sense-check absolute numbers), we can manipulate the above probabilities and re-arrange for the following (unconditional) probabilities of ending at each clinical endpoint:

$$p(\text{F}) = p(\text{F} \mid \text{ICU}^+)\; p(\text{ICU}^+ \mid \text{H}^+)\; p(\text{H}^+)\,, \tag{D.4}$$

$$p(\text{ICU}) = p(\text{ICU}^+ \mid \text{H}^+)\; p(\text{H}^+) - p(\text{F}) \tag{D.5}$$

$$= p(\text{ICU}^+ \mid \text{H}^+)\; p(\text{H}^+)\Big[1 - p(\text{F} \mid \text{ICU}^+)\Big]\,, \tag{D.6}$$

$$p(\text{H}) = p(\text{H}^+) - p(\text{F}) - p(\text{ICU})\,, \tag{D.7}$$

$$p(\text{ED}) = p(\text{ED}^+ \mid \text{SMC})\; p(\text{SMC}) - p(\text{H}^+)\,, \tag{D.8}$$

$$p(\text{GP}) = \Big[1 - p(\text{ED}^+ \mid \text{SMC})\Big]\; p(\text{SMC})\,, \tag{D.9}$$

$$p(\text{nSMC}) = 1 - p(\text{SMC})\;. \tag{D.10}$$

*Appendix D.4. Relative risks*

Infants with risk conditions, including those born premature ($< 37$ weeks) are at higher risk of severe outcomes from RSV infection. We assume all infants born prior to 37 weeks of gestation to be "at-risk", and all other infants "not at-risk". We only consider premature birth as an indicator of risk for infants. Premature births are built into the demographic modelling (see Appendix A), and thus are an easily identifiable at-risk group in the modelling. The prevalence of additional (not premature-birth-correlated) at-risk conditions in infants is not identifiable in available data, to the best of our knowledge.

The probabilities of clinical endpoints in Appendix D.1 are guided by whole-population estimates. That is, numbers such as $N_{\mathrm{H}^+}$ and $N_{\mathrm{ICU}^+}$ include both at-risk and not-at-risk peoples. To stratify these whole-population probabilities into the respective at-risk and not-at-risk probabilities requires us to quantify the relative risk of a given outcome for the at-risk and not-at-risk populations. Explicitly, for a generic outcome "$X$" we wish to split the age-specific whole-population probability $p(X)$ viz.

$$p(X) = p(X \,|\, \text{at-risk})\; p(\text{at-risk}) + p(X \,|\, \text{not-at-risk})\; p(\text{not-at-risk})\,. \tag{D.11}$$

Given the relative risk of outcome $X$, $RR_X = p(X \,|\, \text{at-risk})/p(X \,|\, \text{not-at-risk})$, we can compute $p(X \,|\, \text{at-risk})$ and $p(X \,|\, \text{not-at-risk})$, as we know the prevalence of risk in our model, $p(\text{at-risk})$.

While the provided data does not break down hospitalisations or ICU numbers by premature-birth-status, we access laboratory-confirmed RSV hospital and ICU admission data from the Royal Children's Hospital (RCH), Victoria from 2017–2022 [33].

Data from 2020–2022 from RCH were excluded due to COVID-19 impacts. RCH RSV-coded admission data for under-2-year-olds from 2017–2019 are as follows:

- There are a total of 969 records, of which 266 include ICU admission and 703 have hospitalisation as their final clinical endpoint;
- Of the 266 ICU admissions, 85 are premature births, 181 are not premature births;
- Of the 703 hospitalisations 119 are premature births, 584 are not premature births.

We use these data, combined with Bayes formula, to compute the relative risk for outcome $X$ as

$$RR_X = \frac{p(X \,|\, \text{at-risk})}{p(X \,|\, \text{not-at-risk})}\,, \tag{D.12}$$

$$p(X \,|\, \text{at-risk}) = \frac{p(\text{at-risk} \,|\, X)\; p(X)}{p(\text{at-risk})}\,, \tag{D.13}$$

$$p(X \,|\, \text{not-at-risk}) = \frac{p(\text{not-at-risk} \,|\, X)\; p(X)}{p(\text{not-at-risk})}\,, \tag{D.14}$$

$$\Rightarrow\; RR_X = \frac{p(\text{at-risk} \,|\, X)\; p(\text{not-at-risk})}{p(\text{not-at-risk} \,|\, X)\; p(\text{at-risk})}\,. \tag{D.15}$$

We know $p(\text{not-at-risk})/p(\text{at-risk}) \approx 12.1$ from the proportion of infants that are at-risk (i.e. premature births are 7.61% of the cohort). The remaining terms are estimable as $p(\text{at-risk} \,|\, \mathrm{H}) \approx 119/(584 + 119)$ and

$p(\text{not-at-risk} \mid \text{H}) \approx 584/(584 + 119)$.

Similarly, $p(\text{at-risk} \mid \text{ICU}) \approx 85/(85 + 181)$ and $p(\text{not-at-risk} \mid \text{ICU}) \approx 181/(85 + 181)$. We find $RR_{\text{H}} \approx 2.46$ and $RR_{\text{ICU}} \approx 5.67$.

Expanding the algebra, we combine Equations (D.11) and (D.15) to find

$$p(X \mid \text{not-at-risk}) = \frac{p(X)}{1 + p(\text{at-risk})\ (RR_X - 1)}\,, \tag{D.16}$$

$$p(X \mid \text{at-risk}) = \frac{RR_X\ p(X)}{1 + p(\text{at-risk})\ (RR_X - 1)}\,. \tag{D.17}$$

In summary, we estimate that $RR_{\text{H}} \approx 2.46$ and $RR_{\text{ICU}} \approx 5.69$. We do not know of any data to inform relative risks for other clinical endpoints. However, it is implausible to set them to be one; at-risk infants are likely over-represented at all clinical endpoints (besides nSMC) compared to not-at-risk infants. In the absence of other evidence, we set $RR_{\text{GP}} = RR_{\text{ED}} = RR_{\text{H}}$, and $RR_{\text{F}} = RR_{\text{ICU}}$.

*Appendix D.5. Effect of interventions on clinical outcomes*

Clinical trials show that both mAbs and MV are effective at preventing severe disease outcomes in infants [16–23]. However, the trial endpoints do not map directly to the modelled clinical endpoints described in 2.3. In consultation with ATAGI, we determined fiducial immunisation efficacy inputs for each modelled clinical endpoint, informed by emerging clinical trial evidence, and allowing for a modest increase in efficacy for more severe outcomes. We additionally allow for a slight decrease in the efficacy of each product against each outcome, in at-risk infants [16, 18].

These mappings result in the initial efficacy for each endpoint listed in Table D.5. These initial efficacy values wane over time, according to Equations (B.5) and (B.6) for mAbs and MV respectively. In the model, the complement of the efficacy against each endpoint at the time of an individual's infection is multiplied by their age- and risk-specific probability of that endpoint (see Appendix D.3 and Appendix D.4), reducing the probability of each clinical endpoint.

Table D.5: Initial efficacy against infection and clinical endpoints for mAbs and MV, for both at-risk and not-at-risk infants.

| | | Infection (%) | GP (%) | ED (%) | H (%) | ICU (%) | Death (%) |
|---|---|---|---|---|---|---|---|
| mAbs | Not at-risk | 10 | 50 | 70 | 80 | 85 | 100 |
| | At-risk | 10 | 45 | 65 | 75 | 80 | 100 |
| MV | Not at-risk | 6 | 30 | 50 | 75 | 85 | 100 |
| | At-risk | 6 | 25 | 50 | 70 | 80 | 100 |

### Appendix E. Impact of different seasonal delivery durations and timings

We explore the relative benefits of different seasonal delivery "windows", i.e. combinations of duration of delivery, and when doses start to be delivered to newborns, in Figure E.10. We do this by simulating ten potential delivery windows, and computing the average number of hospitalisations per 100,000 infants that are averted per dose delivered, as compared to the no immunisation scenario. Most scenarios avert a similar number of hospitalisations on average, with the year-round program averting noticeably fewer, as there are more doses delivered off-season, whose efficacy has waned by the onset of the next RSV season.

Delivering doses for only four months between March and June achieves the largest number of hospitalisations averted per dose [2.54 (−0.19, 5.35)], however, there is a large uncertainty due to model stochasticity; the 5th percentile is below zero. The seasonal delivery window of six months between February and July averts 2.33 (0.51, 4.15) hospitalisations per dose, and was decided in consultation with ATAGI as a reasonable middle-ground delivery window to maximise the program impact without compromising the program cost overmuch.

The real-world benefit of different delivery windows of course depends on the timing of the RSV season in each year. In Australia in 2018/2019 hospitalisations in infants began to tick up from February onwards, and began decreasing from August.

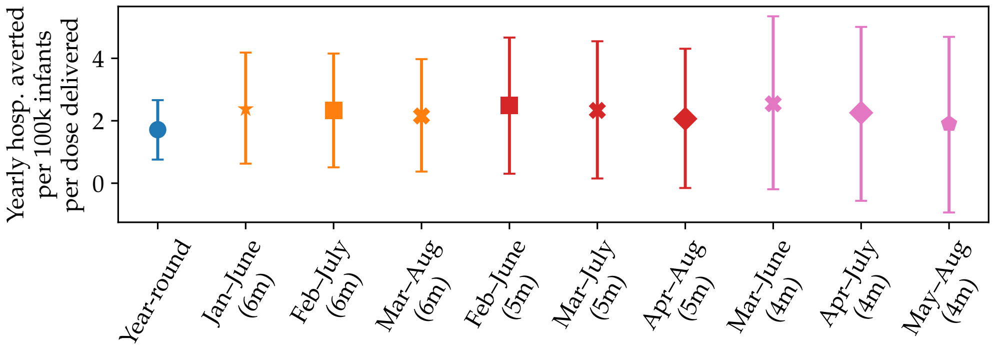


Figure E.10: Mean number (5th–95th percentile) of hospitalisations per 100,000 infants averted per dose delivered, compared to the no immunisation scenario.

### Appendix F. Cost-effectiveness model details

#### *Appendix F.1. Structure of the economic model*

A cohort expected value model is used to evaluate the differences in costs and health outcomes associated with the mAbs and MV compared to no immunisation. Figure F.11 shows the model structure used in the cost-effectiveness analysis. The economic analysis captures only medically attended individuals. For those who are not medically attended, we assume zero costs and no QALY decrement. Medically attended patients may receive treatment in either outpatient settings or inpatient settings. Outpatient settings include GP visits and non-admitted ED visits, and all outpatient cases are assumed to fully recover after receiving treatment. For inpatient infections, cases are classified as either non-ICU or ICU hospitalisations. Inpatient cases either recover or die. The probabilities of infected symptomatic episodes requiring GP visits, non-admitted ED visits, non-ICU hospitalisation, ICU hospitalisation, or resulting in death are specified in the clinical pathways model (see Appendix D). No adverse events following immunisation are included in the economic analysis. This is because the frequencies of adverse events, including grade 3 and serious adverse events, are similar among the interventions compared to placebo, as reported in clinical trials [16, 17, 23].

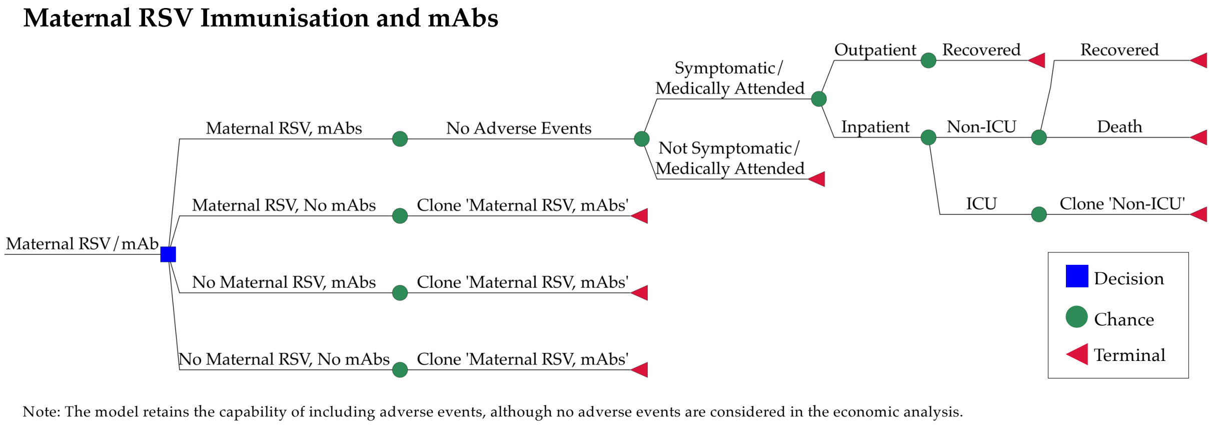


Figure F.11: Structure of the economic model.

*Appendix F.2. Health outcomes*

The outcomes considered in the economic model are: i) the number of GP visit infections; ii) the number of non-admitted ED infections; iii) the number of non-ICU hospitalised infections; iv) the number of ICU hospitalised infections; v) the number of life-years gained; and vi) the number of QALYs gained.

Health related quality of life decrements for each outcome are sourced from Shoukat et al. 2023 [34]. The duration for each health state is based on length of stay data from NHCDC round 26 (2021–2022) when available (see Table F.6). For deaths, we calculate QALY decrements as the total remaining discounted QALY adjusted life expectancy by age of death. We use Australian population norms to reflect a full QALY at a specific age [35]. The source study measured Australian population norms for health-related quality of life using the EQ-5D-5L.

The annual discount rate applied to life years and quality adjusted life expectancy is consistent with the economic model (i.e. 5% in the primary analysis, 0% as the lower bound, and a higher value as the upper bound in the sensitivity analysis). In the sensitivity analysis, the ranges of QALY decrements for each health state are as follows: non-admitted ED presentation ranges from 0.0013 to 0.0035, non-ICU hospitalisation ranges from 0.0018 to 0.0070, and ICU hospitalisation ranges from 0.0122 to 0.0224. These ranges were calculated derived from the length of stay ranges of appropriate AR-DRGs in the NHCDC round 26 (2021–2022) where possible.

*Appendix F.3. Health care resource use and costs*

The costs in the economic analysis include the following:

- Programmatic costs: MV/mAbs dose price and administration cost;

Table F.6: Disutility values and duration of health states used in the economic evaluation. Abbreviations: ABS = Australian Bureau of Statistics; LOS = length of stay; NA = not applicable; NHCDC = National Hospital Cost Data Collection.

| Health state | Mean value | Range | Distribution | Source of estimate |
|---|---|---|---|---|
| **Non-admitted infection** | | | | |
| Disutility | 0.16 | ±50% | Beta | Shoukat et al. 2023 [34] |
| Duration | 5 | 3–8 days | Gamma | Assumption |
| **Non-ICU hospitalisation** | | | | |
| Disutility | 0.41 | ±50% | Beta | Shoukat et al. 2023 [34] |
| Duration | 4.4 | 1.6–6.2 days | Gamma | NHCDC round 26 LOS E62A/B and E70A/B, Brusco 2022 [36] |
| **ICU hospitalisation** | | | | |
| Disutility | 0.60 | ±50% | Beta | Shoukat et al. 2023 [34] |
| Duration | 9.8 | 7.4–13.6 days | Gamma | NHCDC round 26 LOS E41A/B, Brusco 2022[36] |
| **Death** | Age-specific, based on discounted life expectancy | NA | NA | Australian population norms for health-related quality of life are sourced from Redwood et al. 2024 [35] and Australian population life expectancy is sourced from the ABS[1]. |

- Outpatient care: cost of GP visits, cost of non-admitted ED visits, and cost of RSV viral tests; and
- Inpatient care: cost of non-ICU hospitalisation and cost of ICU hospitalisation.

Table F.7 presents the unit costs, range, and distribution assumptions applied in the economic model.

*Appendix F.3.1. Programmatic costs*

Immunisation program costs include the MV/mAbs dose price, wastage and MV/mAbs administration. Australian MV/mAbs dose prices are not currently available. In the base case analysis, assumed dose prices were based on the U.S. Centers for Disease Control and Prevention prices ($US 230.09 for Abrysvo™ and $US 414.75 for Beyfortus™), which were converted to AUD assuming an exchange rate of 0.62964 (AUD to USD). Dose prices were then moderated down by 43.6% based on reported average differences in medicines prices between the United States and Australia [39]. A range of product prices is explored in sensitivity analyses. For the upper estimate, we used U.S. prices without reduction. For the lower estimate, we used A$75 for MV based on a cost-effectiveness analysis from Australia [40]. The primary analysis estimate for mAbs was also used as the lower estimate given that we did not identify any published studies or public procurement prices lower than this estimate.

Due to the lack of available data on MV/mAbs wastage rates in Australia, we assume a wastage rate of 5% based on maximum target rate of wastage [37]. In the sensitivity analysis, we consider a range of 0% to 10%.

The cost of MV/mAbs administration is based on the weighted average cost of Medicare Benefits Schedule (MBS) item 23 (A$42.85) and item 82205 (A$31.05), as per consultation with the RSV ATAGI subgroup.

We assume that 50% of doses are administered by a GP and 50% by a nurse. A range of administration costs (from A$31.05 to A$42.85) is explored in sensitivity analysis.

Programmatic costs related to MV/mAbs roll-out, delivery, and storage are not considered. Similarly, costs associated with surveillance for effectiveness or safety are excluded.

*Appendix F.3.2. Outpatient care costs*

For non-admitted cases requiring GP visits, we assume that patients will incur costs for two GP consultations (A$42.85, MBS item 23) and one RSV test (A$28.65, MBS item 69494). We do not include radiology costs because it is not routinely used for diagnosing RSV cases in outpatient settings [41, 42].

The cost of a non-admitted ED visit (A$724) is sourced from the national average cost of non-admitted ED attendances reported in NHCDC round 26 (2021–2022).

A range of unit costs of outpatient services (±50%) is explored in sensitivity analyses.

*Appendix F.3.3. Inpatient care costs*

We assume that all RSV-related hospitalisations present to ED first and then are admitted to hospital. The admitted ED visit per case (A$1335) is based on the national average cost of admitted ED attendances reported in NHCDC round 26 (2021–2022).

For non-ICU hospitalisations, the following NHCDC AR-DRG codes are considered: E62A (Respiratory Infections and Inflammations, Major Complexity), E62B (Respiratory Infections and Inflammations, Minor Complexity), E70A (Whooping Cough and Acute Bronchiolitis, Major Complexity) and E70B (Whooping Cough and Acute Bronchiolitis, Minor Complexity).

For ICU hospitalisations, the following NHCDC AR-DRG codes are considered: E41A (Respiratory System Disorders W Non-Invasive Ventilation, Major Complexity) and E41B (Respiratory System Disorders W Non-Invasive Ventilation, Minor Complexity).

The weighted average DRG costs per non-ICU hospitalisation (A$7,980) and per ICU hospitalisation (A$25,490) are calculated based on the number of episodes. The selection of AR-DRG codes is informed by a study of the annual cost burden of children with RSV in Australia [36] and prior PBAC submissions [43]. All inpatient care costs are varied by ±50% in the sensitivity analyses.

Table F.7: Health care resource items and unit costs included in the economic evaluation.

| Resource item | Primary analysis value | Range | Distribution | Source |
|---|---|---|---|---|
| **Dose and administration costs** | | | | |
| Cost per mAbs dose | A$290 | A$290–A$660 | Uniform | CDC pricing scaled down by 43.6% |
| Cost per MV dose | A$160 | A$75–A$370 | Uniform | CDC pricing scaled down by 43.6% |
| Cost per dose administration (newborn & maternal) | A$36.35 | A$31.05–A$42.85 | Gamma | MBS item 23 & MBS item 82205 |
| **Outpatient costs** | | | | |
| Cost of GP consultation for RSV-related reasons | A$41.40 | ±50% | Gamma | MBS item 23 |
| Number of visits per case | 2 | 1–3 | Discrete | Assumption |
| Cost of non-admitted ED visit | A$724 | ±50% | Gamma | NHCDC round 26 (2021–2022) (national average cost of non-admitted ED) |
| Cost of virologic tests | A$28.65 | ±50% | Gamma | MBS item 69494 |
| **Hospitalisation costs** | | | | |
| Cost of admitted ED visit | A$1,335 | ±50% | Gamma | NHCDC round 26 (2021–2022) (national average cost of admitted ED) |
| Cost of non-ICU hospitalised infection | A$7,980 | ±50% | Gamma | Average E62A, E62B, E70A and E70B costs weighted by the number of episodes from NHCDC round 26 (2021–2022) |
| Cost of ICU hospitalised infection | A$25,490 | ±50% | Gamma | Average E41A and E41B costs weighted by the number of episodes from NHCDC round 26 (2021–2022) |
| **Other items** | | | | |
| mAb/MV wastage rate | 5% | 0%–10% | Beta | Assumed based on [37]. |
| Annual discounting rate | 5% | 0%–5% | Fixed at 5% in the probablistic sensitivity analysis | Based on national guidelines [38]. |

CDC = U.S. Centers for Disease Control and Prevention; ED = emergency department; ICU = intensive care unit; MBS = Medicare Benefits Schedule; NHCDC = National Hospital Cost Data Collection.

### Appendix G. Results for mAbs-only and MV-only scenarios

Figure G.12 shows the average modelled annual incidence of RSV infections, GP and ED visits, hospitalisations, ICU admissions, and deaths in infants using only mAbs.

The numbers of clinical endpoints averted, with average doses delivered under each scenario, are tabulated in Table G.8. The best scenario in preventing the most severe clinical outcomes is when all infants are given mAbs at birth, as expected. The next-best is when either 70% of infants are given mAbs at birth year-round, or mAbs are given at birth to 100% of infants born Feb through July (i.e. seasonal administration). The latter scenario requires 40% fewer doses than the former. While the risk-targeted scenario prevents fewer total clinical outcomes, considerably fewer doses are delivered in this scenario.

Figure G.13 and Table G.9 show the cost-effectiveness results for the mAbs-only scenarios, using the mean values across all clinical pathways outputs from the transmission model. Based on modelled dose price (mAbs = A$290), the results indicate that immunising 100% of at-risk newborns with mAbs year-around or seasonal immunisation of all newborns is cost-saving, while other mAbs-only immunisation scenarios have high ICER values (>200,000/QALY gained).

Table G.8: Average modelled numbers (5th–95th percentile) of clinical endpoints of RSV infection per year in infants scaled to the size of the Australian population in 2018/2019, in a no intervention scenario. Also tabulated are the average doses delivered per year and numbers of clinical endpoints that are averted per year under different mAbs-only immunisation scenarios.

| **Scenario** | | **GP ('000)** | **ED ('000)** | **H ('000)** | **ICU ('000)** | **Deaths** |
|---|---|---|---|---|---|---|
| No intervention | | 66.0 (61–70) | 15.4 (13–18) | 16.0 (14–18) | 0.7 (0–1) | 2.8 (0–9) |
| | **mAbs delivered ('000)** | **GP averted ('000)** | **ED averted ('000)** | **H averted ('000)** | **ICU averted ('000)** | **Deaths averted** |
| 100% mAbs year-round | 330 | 15.2 (10–20) | 7.4 (5–10) | 6.8 (5–9) | 0.4 (0–1) | 2.0 (0–6) |
| 100% at-risk mAbs year-round | 26 | 2.4 (−2–6) | 1.2 (−1–4) | 1.1 (−1–3) | 0.1 (0–1) | 0.6 (0–2) |
| 100% mAbs, seasonal | 163 | 10.2 (5–15) | 5.2 (3–8) | 4.6 (3–7) | 0.3 (0–1) | 1.4 (0–4) |
| 70% mAbs year-round | 231 | 10.9 (6–16) | 5.2 (3–8) | 4.8 (3–7) | 0.3 (0–1) | 1.5 (0–4) |
| 50% mAbs year-round | 165 | 7.9 (3–13) | 3.9 (1–7) | 3.5 (2–6) | 0.2 (0–1) | 1.1 (0–3) |

ED = emergency department presentations; GP = general practitioner visits; H = hospitalisations; ICU = intensive care unit admissions.

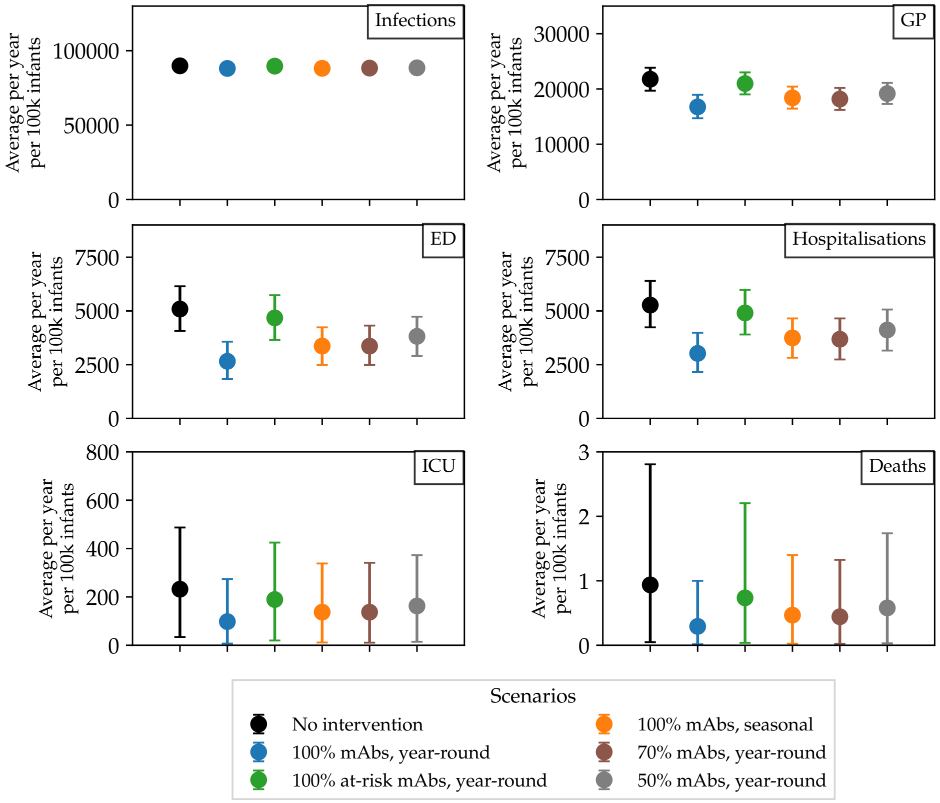


Figure G.12: Mean incidence (5th–95th percentile) of medically-attended RSV (per 100,000 infants <1 year of age) that leads to GP, ED visits, hospitalisations, ICU admissions and deaths in scenarios using only mAbs.

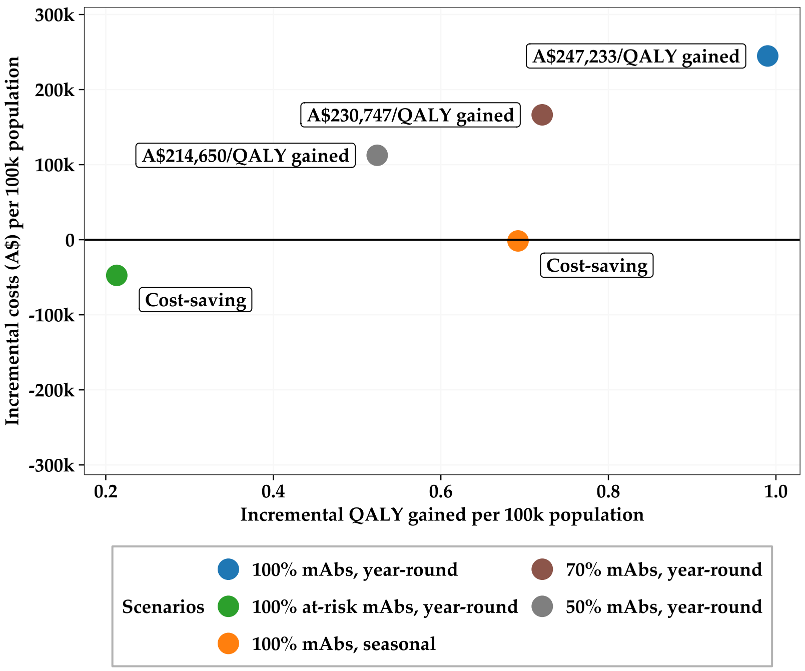


Figure G.13: Mean incremental costs and incremental QALY gained per 100,000 population in mAbs-only scenarios compared to no intervention.

Table G.9: Mean costs, quality-adjusted life years gained, and incremental cost-effectiveness ratios, per 100,000 population (mAbs-only scenarios)

| **Scenario** | **Total Cost** | **QALY decrements** | **Incr. Cost** | **Incr. QALY** | **ICER (/QALY gained)** |
|---|---|---|---|---|---|
| No intervention | A$1,443,902 | −2.52 | Reference | Reference | Reference |
| 100% mAbs year-round | A$1,688,712 | −1.53 | A$244,810 | 0.99 | A$247,233 |
| 100% at-risk mAbs year-round | A$1,396,412 | −2.30 | −A$47,490 | 0.21 | Cost-saving |
| 100% mAbs seasonal | A$1,442,309 | −1.82 | −A$1,593 | 0.69 | Cost-saving |
| 70% mAbs year-round | A$1,610,266 | −1.79 | A$166,364 | 0.72 | A$230,747 |
| 50% mAbs year-round | A$1,556,381 | −1.99 | A$112,479 | 0.52 | A$214,650 |

ICER = incremental cost-effectiveness ratio; QALY = quality-adjusted life-year.

The MV-only scenarios were explored to evaluate the potential impact of different MV programs that vary by coverage. MV doses were delivered year-round to pregnant people between 28 and 36 weeks gestation. Seasonal MV programs were not explored per expert advice regarding the logistical feasibility and equity of such potential programs.

Figure G.14 shows the average modelled annual incidence of RSV infections, GP and ED visits, hospitalisations, ICU admissions, and deaths in infants using only MV.

The numbers of clinical endpoints averted, with average doses delivered under each scenario, are tabulated in Table G.10. The best MV-only scenario in preventing clinical outcomes is when all pregnant people (100% coverage) are given MV between 28–36 weeks gestation. As expected, as coverage reduces to 'feasible' levels (70% and 50%) fewer clinical outcomes are prevented, however, there is still a noticeable impact on disease compared to the 'no intervention' scenario.

Table G.10: Average modelled numbers (5th–95th percentile) of clinical endpoints per year of RSV infection in infants scaled to the size of the Australian population in 2018/2019, in a no intervention scenario. Also tabulated are the average doses delivered per year and numbers of clinical endpoints that are averted per year under different MV-only immunisation scenarios.

| **Scenario** | | **GP ('000)** | **ED ('000)** | **H ('000)** | **ICU ('000)** | **Deaths** |
|---|---|---|---|---|---|---|
| No intervention | | 66.0 (61–70) | 15.4 (13–18) | 16.0 (14–18) | 0.7 (0–1) | 2.8 (0–9) |
| | **MV delivered ('000)** | **GP averted ('000)** | **ED averted ('000)** | **H averted ('000)** | **ICU averted ('000)** | **Deaths averted** |
| MV 100% year-round | 324 | 11.3 (6–16) | 5.6 (3–8) | 7.4 (5–10) | 0.4 (0–1) | 1.9 (0–6) |
| MV 70% year-round | 227 | 7.7 (3–13) | 3.9 (1–7) | 5.1 (3–7) | 0.3 (0–1) | 1.4 (0–4) |
| MV 50% year-round | 162 | 5.5 (1–10) | 2.8 (0–5) | 3.6 (2–6) | 0.2 (0–1) | 1.0 (0–3) |

ED = emergency department presentations; GP = general practitioner visits; H = hospitalisations; ICU = intensive care unit admissions; MV = maternal RSV vaccination.

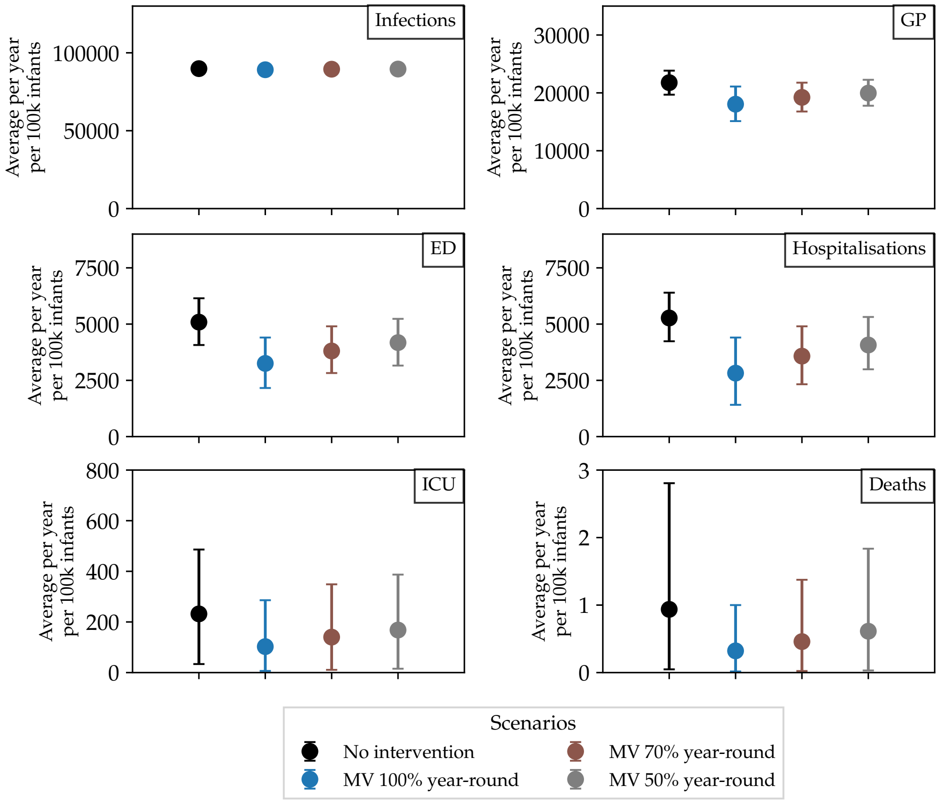


Figure G.14: Average incidence (5th–95th percentile) of medically-attended RSV (per 100,000 infants <1 year of age) that leads to GP, ED visits, hospitalisations, ICU admissions and deaths in scenarios using only MV.

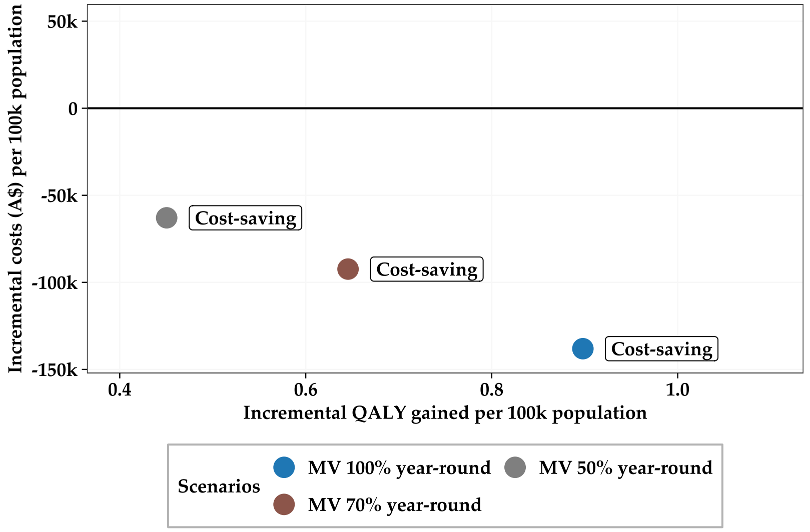


Figure G.15: Mean incremental costs and incremental QALY gained per 100,000 population in MV-only scenarios compared to no intervention.

Figure G.15 and Table G.11 show the cost-effectiveness results for the MV-only scenarios. Based on modelled dose price (MV = $160), the results indicate that all MV-only scenarios are cost-saving, with the 100% year-round program achieving the highest QALY gains, while also saving the most health system costs.

Table G.11: Mean costs, quality-adjusted life years gained, and incremental cost-effectiveness ratios, per 100,000 population (MV-only scenarios)

| **Scenario** | **Total Cost** | **QALY decrements** | **Incr. Cost** | **Incr. QALY** | **ICER (/QALY gained)** |
|---|---|---|---|---|---|
| No intervention | A$1,443,902 | −2.52 | Reference | Reference | Reference |
| MV 100% year-round | A$1,305,823 | −1.62 | −A$138,079 | 0.90 | Cost-saving |
| MV 70% year-round | A$1,351,536 | −1.87 | −A$92,367 | 0.65 | Cost-saving |
| MV 50% year-round | A$1,380,983 | −2.06 | −A$62,920 | 0.45 | Cost-saving |

ICER = incremental cost-effectiveness ratio; MV = maternal RSV vaccination; QALY = qualty-adjusted life-year.

For all immunisation programs, changes in coverage (i.e. from 100%, 70%, to 50%) are unlikely to alter cost-effectiveness estimates. We also observe that at the same coverage levels (e.g. 100% mAbs year-round and MV 100% year-around), the mAbs-only programs result in slightly higher incremental QALYs gained compared to the MV-only programs.

### Appendix H. Sensitivity analyses

Many of our model parameters are uncertain. We tabulate in Table H.12 the parameters that were varied as part of our sensitivity analyses, along with the ranges and sampling distributions. For the probabilistic sensitivity analyses, not all parameters are sampled uniformly within the range specified. In particular, the multiplier on the incidence of each medically attended outcome is drawn from a triangular distribution, rising from 0.9 to a mode at 1, then dropping from 1 to 2. We choose this distribution and range as we believe that the observed RSV-coded infant clinical burden in Australian hospitalisation data are more likely an under-estimate than an over-estimate, although we allow for both possibilities.

We conducted a one-way sensitivity analysis for the combined scenario in which 70% of pregnant people are given MV year-round, with 50% of newborns born without protection from MV given mAbs year-round program (Figure H.16). ICERs were most sensitive to dose price (particularly MV) and the cost per case of non-ICU hospitalisation infection, with the scenario no longer being cost saving at the most conservative estimates for these parameters. ICERs were also sensitive to the incidence of hospitalisation, however, the scenario was still cost-saving when when we assume a 10% over-ascertainment of RSV-coded hospitalisations (represented as a 0.9 multiplier applied to aggregated data).

Table H.12: List of parameters varied as part of the sensitivity analyses, along with the ranges within which they are varied.

| **Parameter** | **Range**[a] | **Distribution**[b] | **Notes** |
|---|---|---|---|
| Multiplier on incidence of each medically attended outcome | 0.9–2 | Triangular | Assumption[c] |
| mAbs immunisation efficacy | ±20% | Uniform | Assumption |
| MV immunisation efficacy | ±20% | Uniform | Assumption |
| Cost per mAbs dose | A$290–A$660 | Uniform | Upper: United States pricing [44]; Lower: same as base case |
| Cost per MV dose | A$75–A$370 | Uniform | Upper: United States pricing [44]; Lower: Nazareno et al. 2025 [40] |
| Cost per dose administration | ∼ ±15% | Gamma | Upper: MBS item 23; Lower: MBS item 82205 |
| Cost of each medically attended outcome | ±50% | Gamma | Assumption[d] |
| Number of GP visits per case | 1–3 | Discrete | Assumption |
| Cost of virologic tests | ±50% | Gamma | Assumption |
| Annual discounting rate | 0%–5% | Fixed at 5%[e] | Based on national guidelines [38] |
| Wastage rate | 0%–10% | Beta | Assumption |
| Disutility of each medically attended outcome | ±50% | Beta | Assumption. |
| Duration of non-admitted infection | 3–8 days | Gamma | See note[f] |
| Duration of non-ICU hospitalisation | 1.6–6.2 days | Gamma | See note[f] |
| Duration of ICU hospitalisation | 7.4–13.6 days | Gamma | See note[f] |

[a] Range used as upper and lower bounds for deterministic sensitivity analyses and confidence intervals for probabilistic sensitivity analysis.
[b] Distribution used for probabilistic sensitivity analysis.
[c] Incidence in the primary analysis is based on the Australian hospitalisation dataset of RSV-coded cases, which we believe is more likely to under-estimate than over-estimate infant clinical burden.
[d] Range encompasses the highest and lowest costs from the mix of DRG codes informing the primary analysis.
[e] Parameter was not varied in probabilistic sensitivity analysis.
[f] Range encompasses the highest and lowest length of stay from the mix of DRG codes informing the primary analysis.

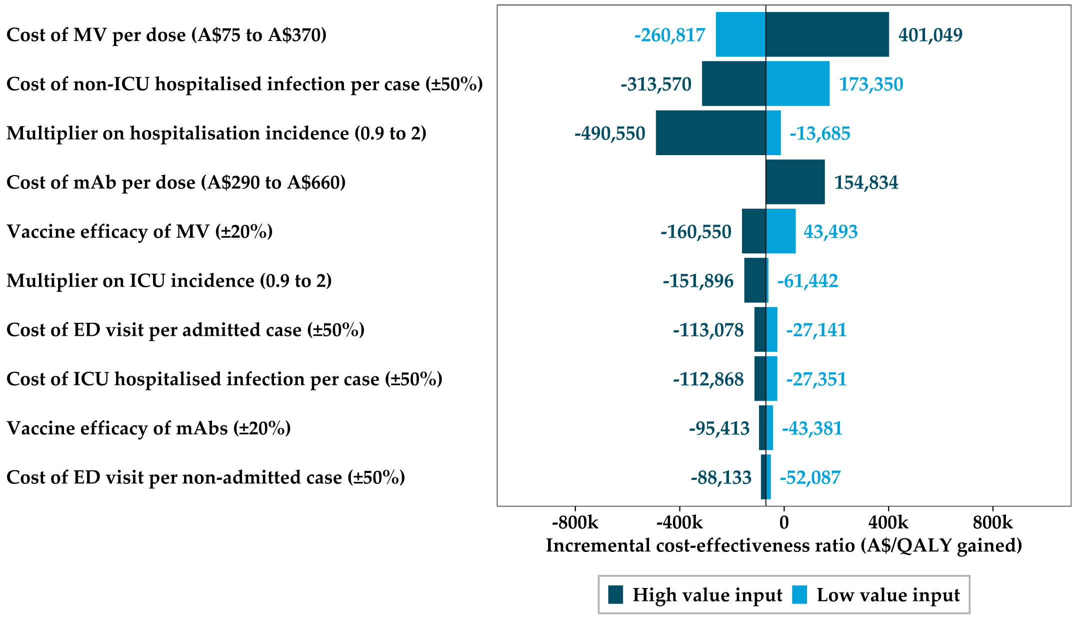


Figure H.16: One-way sensitivity analysis showing the incremental cost-effectiveness ratio (A$/QALY gained) for the scenario in which MV is delivered to 70% of pregnant people year-round, with mAbs delivered year-round to 50% of newborns born without protection from MV.
